# Contemporary Burden of Cardiovascular Disease in Pregnancy: Insights from a Real-World Pregnancy Electronic Health Record Cohort

**DOI:** 10.1101/2025.01.28.25320930

**Authors:** Emily S. Lau, Valentina D’Souza, Yunong Zhao, Christopher Reeder, Rachel Goldberg, Katherine E. Economy, Mahnaz Maddah, Shaan Khurshid, Patrick T. Ellinor, Jennifer E. Ho

## Abstract

**Importance:** Cardiovascular disease (CVD) is the leading cause of maternal morbidity and mortality, however the contemporary burden and secular trends in pregnancy-related CV complications are not well characterized.

**Objective:** We sought to examine contemporary trends in prevalence of maternal cardiometabolic comorbidities and established CVD, as well as future pregnancy-related CV complications across a large multi-institutional health system.

**Design:** Retrospective analysis of longitudinal electronic health record (EHR)-based cohort of pregnancies

**Setting:** Multi-institutional healthcare network in New England

**Participants:** Pregnancy encounters between 2001 to 2019 identified using diagnosis and procedure codes followed by manual adjudication within a previously validated primary care EHR cohort. Estimated gestational ages recovered from unstructured notes using regular expressions (RegEx) were used to define individual pregnancy episodes.

**Main Outcomes and Measures:** We quantified the prevalence of maternal cardiometabolic comorbidities and established CVD at time of pregnancy, as well as the incidence of pregnancy-related CV complications assessed within 1 year postpartum. We examined trends in cardiometabolic risk factors and CVD burden over nearly two decades.

**Results:** Our EHR pregnancy cohort comprised 57,683 pregnancies among 38,997 individuals (mean age range at start of pregnancy 27 to 37 years). RegEx recovered gestational age for 74% of pregnancies, with good correlation between gestational age ascertained via RegEx vs manual review (Pearson r 0.9). Overall prevalence of maternal CVD was 4% (age-adjusted 7%) and increased over 19 years of follow-up (age-adjusted prevalence of maternal CVD: 1% in 2001 to 7% in 2019, p <0.001). The incidence of pregnancy-related CV complications was 15% (age-adjusted 17%) and also increased over the follow-up period (age-adjusted incidence 11% in 2001 to 14% in 2019, p <0.001). Finally, CV complications were more likely to occur in individuals with greater burden of maternal CV comorbidities and CVD (diabetes: 6% vs 3%, hypertension: 23% vs 5%, pre-existing CVD: 10% vs 3%, P<0.001 for all).

**Conclusions and Relevance:** Analysis of a large-scale EHR-based pregnancy cohort spanning two decades demonstrates rising prevalence of both maternal cardiometabolic comorbidities and CVD at the time of pregnancy, as well as increasing incidence of subsequent pregnancy-related CV complications. Pregnancy represents a critical opportunity for cardiometabolic health optimization.

**KEY POINTS:** *Question:* What are the contemporary real-world trends in the prevalence of maternal cardiovascular comorbidities and cardiovascular disease and incidence of cardiovascular complications in pregnancy?

*Findings:* In an analysis of 57,683 pregnancies among 38,997 individuals from a large scale EHR-based pregnancy cohort, prevalence of maternal cardiometabolic comorbidities and cardiovascular disease and incidence of pregnancy-related cardiovascular complications increased over the course of nearly two decades.

*Meaning:* The contemporary burden of pregnancy-related cardiovascular complications is rising at an alarming rate and highlights pregnancy as a critical opportunity for cardiovascular health optimization.

## INTRODUCTION

The United States has the highest maternal mortality rate among industrialized countries, with an estimated rate of 32.9 deaths/100,000 live births in 2021. Moreover, maternal deaths are rising at an alarming rate, particularly among non-Hispanic Black individuals.^1–5^ Cardiovascular diseases (CVD) are the leading cause of maternal morbidity and mortality, and account for over one-third of maternal deaths.^3,6,7^ Pregnant individuals with pre-existing CVD experience greater maternal morbidity and mortality than individuals without CVD due to greater risk of cardiovascular (CV) complications during pregnancy.^5,8–10^

There is therefore an urgent need to better understand the contemporary burden of CVD during pregnancy to enable identification of individuals at high risk for developing pregnancy-related CV complications. While existing studies have documented prevalence of maternal CVD in pregnancy, data on the incidence and trends in CV complications during pregnancy, particularly among individuals without a history of CVD, are lacking. This is due in part to the paucity of large pregnancy cohorts with robust cardiovascular outcome data. Most pregnancy cohorts with detailed outcome data are small (∼500-1000 patients) and largely restricted to individuals with existing CVD, while large scale cohorts derived from nationwide databases are limited by misclassification of outcomes and absence of granular clinical data.^11–13^ To address these limitations, we developed a multi-institutional electronic health record (EHR)-based cohort of >57,000 pregnancies originating from a previously validated primary care cohort with rich clinical data and rigorously defined longitudinal CV outcomes (**Figure 1**). Leveraging this novel cohort, we sought to describe secular trends in prevalent maternal cardiometabolic comorbidities and established CVD at time of pregnancy, and development of subsequent CV complications during pregnancy.

**Figure 1.**
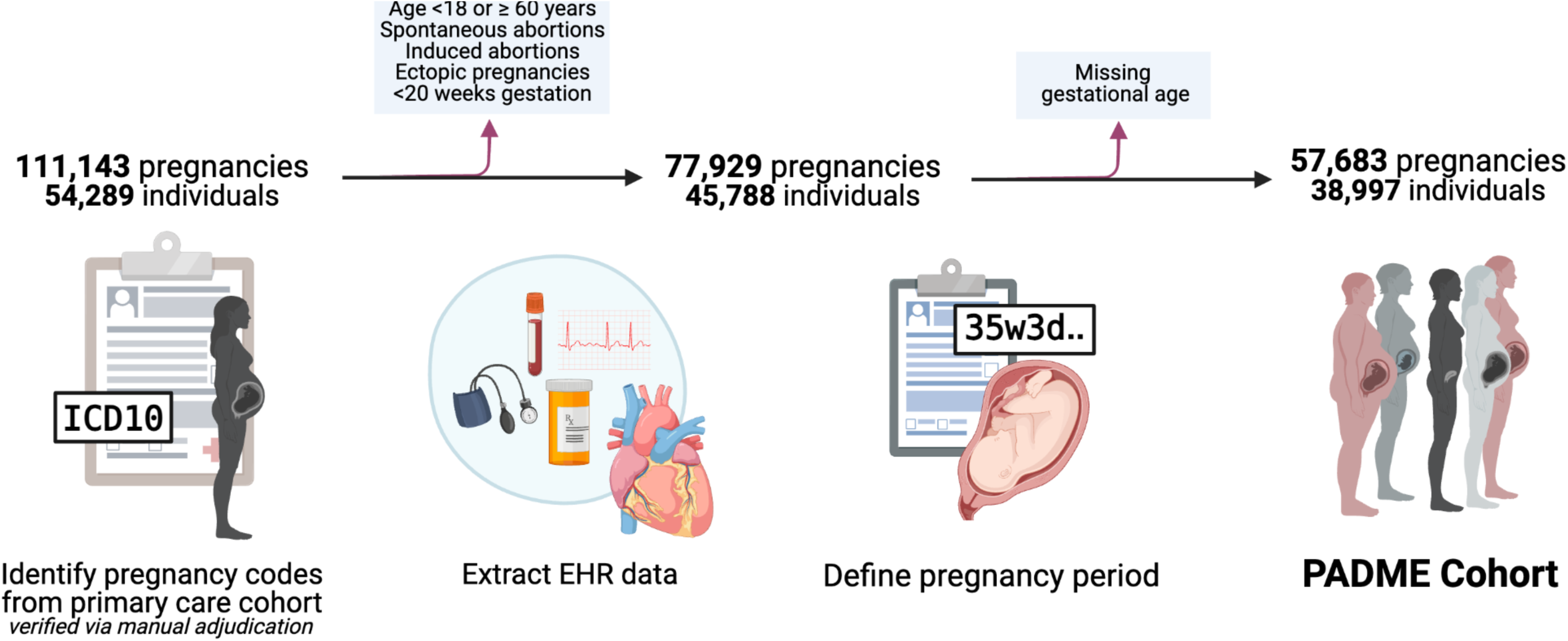
Overview of PADME Construction. Displayed is a graphical overview of the construction of the Predictive Analysis with Deep Learning Models for Maternal Endpoints (PADME) Cohort. PADME comprises electronic health record (EHR) data for 57,683 pregnancies from 38,997 individuals. Pregnancies were identified from a validated primary care EHR cohort receiving longitudinal ambulatory care based on presence of pregnancy endpoint diagnostic codes. PADME is an indexed file system that contains diverse protected health information-minimized data types including vital signs, billing codes, narrative nodes, medications, laboratory tests, and diagnostic tests. Abbreviations: EHR = electronic health record, ICD = International Classification of Disease, PADME = Predictive Analysis with Deep Learning Models for Maternal Endpoints.

## METHODS

### Cohort construction

We developed the Predictive Analysis with Deep Learning Models for Maternal Endpoints (PADME) cohort, an EHR-based pregnancy cohort in Mass General Brigham (MGB), a multi-institutional healthcare network spanning eleven tertiary care and community hospitals in New England. Pregnancy encounters at MGB were identified from a validated primary care EHR cohort of 520,868 individuals receiving longitudinal primary care between 2001 to 2019 using International Classification of Disease, 9th and 10th revision (ICD-9 and 10) diagnosis and Current Procedural Terminology (CPT) codes (definitions in **eTable 1**).^14,15^ The MGB Institutional Review Board approved study protocols.

From a total sample of 111,143 candidate pregnancies (among 54,289 women), we excluded pregnancies from individuals aged <18 or ≥ 60 years of age (n=1,256), spontaneous abortions (n=19,511), induced abortions (n=8103), ectopic pregnancies (n = 1032), pregnancies < 20 weeks gestation (n = 3312), and those with missing gestational age (n=20,246), yielding a final cohort of n=57,683 pregnancies (among 38,997 individuals).

### Cohort validation

We validated the PADME cohort construction algorithm via manual adjudication of the EHR. Two independent physicians reviewed 200 randomly selected candidate pregnancy encounters to verify pregnancy status. To assess interobserver agreement, 20 pregnancy encounters were overlapping between the two reviewers. Manual adjudication demonstrated a positive predictive value (PPV) of 91% (95% CI 86% to 95%) and inter-observer agreement of 100% (95% CI 83% to 100%). Pregnancy encounters met a pre-specified PPV ≥ 85% to proceed with cohort construction.

### Using regular expressions to ascertain gravidity, parity, and gestational age from unstructured clinical notes

As data on gravidity, parity, and estimated gestational age are not routinely captured in structured data fields or by diagnostic codes, we employed regular expressions (RegEx) to ascertain these data at time of delivery for each pregnancy. Documentation of gravidity, parity, and gestational age in clinical notes typically follow a standardized format (e.g. G*n*P*n* or gravida *n*, para *n*). We crafted RegEx to identify these patterns and retrieve these data from clinical notes (**eMethods**). **eFigure 1** demonstrates an example of a clinical note with RegEx-extracted gravidity, parity, and gestational age data. We validated the RegEx approach via manual review of 200 randomly selected pregnancy encounters in the EHR by two reviewers.

### Defining pregnancy episodes

We defined the pregnancy episode for each individual pregnancy in the PADME cohort as the time period between the date of the estimated last menstrual period (gestational age 0) to the date of delivery. Further details including the stepwise approach to constructing each pregnancy episode are summarized in **eMethods** and **eFigure 2**. We validated this approach via manual EHR review of 200 randomly selected pregnancy encounters.

**Figure 2.**
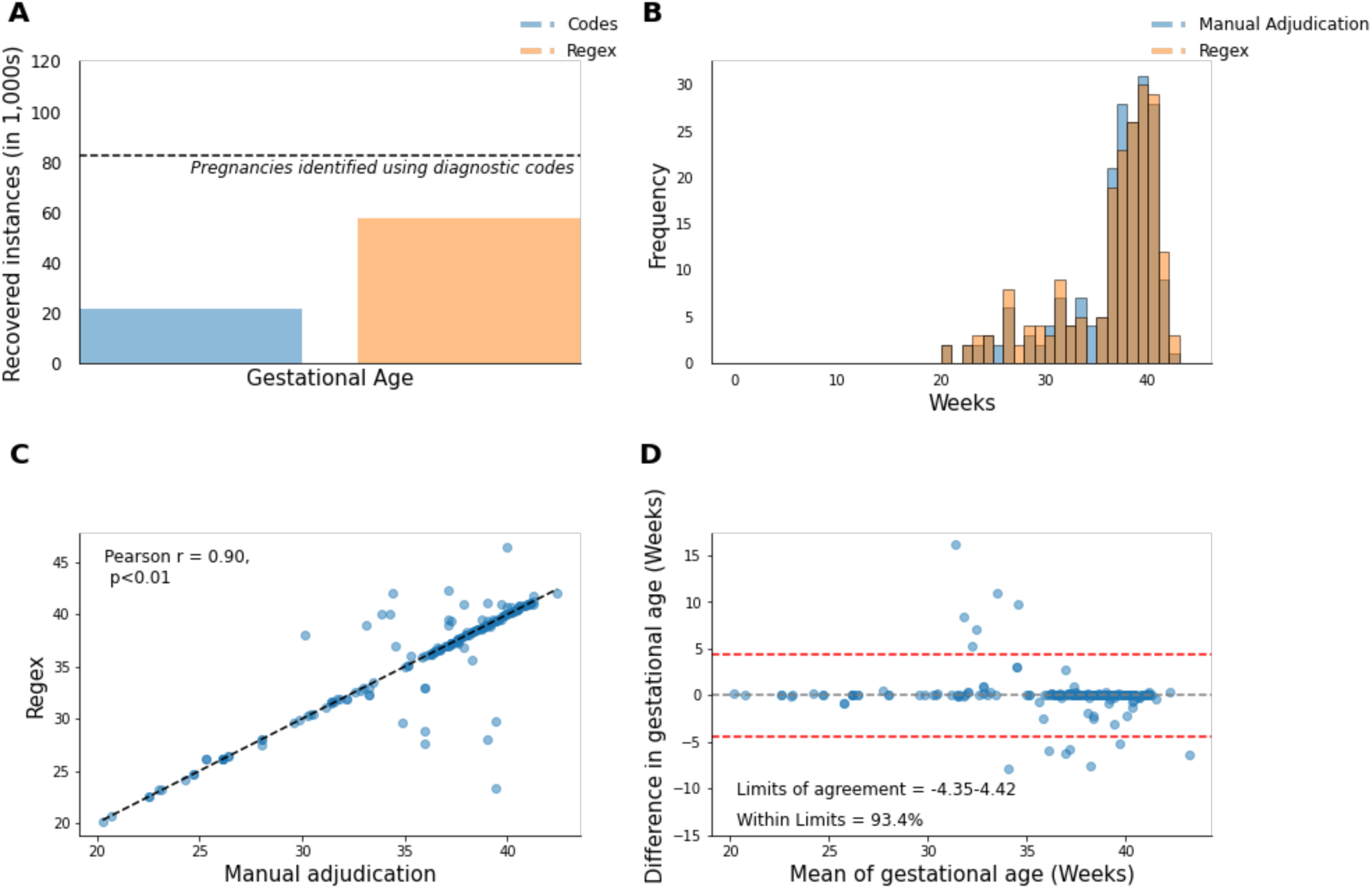
Recovery of estimated gestational age with RegEx. Displayed is a summary of the yield of our approach to extract estimated gestational age data for pregnancies in PADME using RegEx. **Panel A** depicts the total number of pregnancies with available gestational age data using diagnostic codes (denoted in blue) and after recovery with RegEx (denoted in yellow). The dashed line indicates the total number of pregnancies identified using pregnancy delivery codes. **Panels B-D** display the agreement between gestational age obtained from diagnostic codes and those obtained using RegEx. **Panel B** displays the distribution of values obtained from diagnostic codes (blue), RegEx (yellow), and manual adjudication (red). **Panel C** shows the correlation between code values (x-axis) and RegEx values (y-axis). **Panel D** displays a Bland-Altman plot assessing agreement between paired code vs RegEx values for gestational age. The x-axis shows the mean of the paired values and the y-axis displays the difference between the paired values. Positive values indicate code values greater than the corresponding RegEx values and negative values indicate RegEx values greater than corresponding code values. The black dashed line depicts the overall mean difference, and the red dashed line depicts the estimated 95% limits of agreement. Abbreviations: RegEx = regular expression.

### Assessment of baseline clinical variables

Clinical exposures including demographic data, vital signs, prevalent comorbidities and disease, smoking status, and alcohol use were assessed from one year preceding the start of the pregnancy episode (gestational age 0) to the end of the first trimester of pregnancy (gestational age 13 weeks) except for smoking status and alcohol use (assessment periods described in **eMethods**). Clinical covariates definitions are detailed in **eMethods** and **eTables 2-3**.

### Pregnancy-related cardiovascular complications

Pregnancy-related CV complication was defined as the composite of maternal death, major adverse CV events (MACE), and hypertensive disorders of pregnancy (HDP) assessed from the end of the first trimester (gestational age 13 weeks) to 1 year postpartum as typically defined.^16^

Maternal death was ascertained from the Social Security Death Index or MGB internal documentation of death.

MACE included myocardial infarction (MI), heart failure (HF), vascular dissection, thromboembolism (venous, pulmonary and systemic), cerebrovascular disease (transient ischemic attack [TIA] and hemorrhagic and ischemic cerebrovascular attack), ventricular and atrial arrhythmias, and cardiac arrest. Clinical definitions for MI and HF were previously validated with PPV ≥85%, while remaining MACE events were defined using ICD-9 or ICD-10 code groupings (**eTable 2**).^15^

HDP, including chronic hypertension, gestational hypertension, preeclampsia with and without severe features, superimposed preeclampsia, eclampsia, and hemolysis, elevated liver enzymes, low platelets (HELLP) syndrome, were identified using ICD-9 and ICD-10 codes (**eTable 3**). HDP outcomes were validated by manual review of 200 medical records by two independent physicians with PPV of 85% (95% CI 79% to 90%) and inter-observer agreement of 95% (95% CI 91% to 98%).

### Statistical analysis

Clinical and pregnancy characteristics were summarized using Student’s t-test, chi-square test, or Wilcoxon rank sum test as appropriate. For estimated gestational age, we manually reviewed the EHR and compared agreement with gestational age ascertained via our RegEx approach vs diagnostic codes using Pearson correlations and Bland-Altman plots.

We first quantified the prevalence of maternal cardiometabolic comorbidities and existing CVD over the entire cohort follow-up time and for the following time increments (2001-2005, 2006-2010, 2011-2015, 2016-2019). We next calculated the incidence of pregnancy-related CV complications as the number of incident events divided by the total number of pregnancies during the specified time period. Incidence rates were stratified by maternal age at time of delivery (<25 years, 26-35 years, 36-45 years, >45 years). Prevalence and incidence rates were adjusted for maternal age using the direct method with weights derived from the 2010 U.S. Census Data.^17^ Finally, we examined secular trends in age-adjusted prevalence of maternal cardiometabolic risk factors and existing CVD and incidence of CV complications in pregnancy using age-and year-adjusted Poisson regression models to estimate incidence rate ratio over time (compared with 2001-2005). Sensitivity analyses examined trends in age-adjusted prevalence and incidence restricted to first pregnancies available in PADME.

Analyses were performed in R version 4.4.0 and Python version 3.^18^ Two-sided p-values <0.05 were considered significant.

## RESULTS

### PADME Cohort

The PADME cohort comprised 57,683 pregnancies among 38,997 individuals (mean age range at start of pregnancy 27 to 37 years). **Table 1** summarizes pregnancy-level demographic and clinical characteristics (**eTable 4** displays individual-level characteristics). Among 57,683 total pregnancies, 52,551 (91%) were classified as live births, 38 (0.1%) mixed births (multiple gestations with live and non-live births), 515 (1%) stillbirth, and 4579 (8%) unclassified deliveries (**eTable 5**).

**Table 1.** Baseline demographics and clinical characteristics of pregnancies included in PADME.

|  | **Total**<br>**PADME** | **Cardiovascular**<br>**complication** | **Without**<br>**cardiovascular**<br>**complication** | **p-value** |
| --- | --- | --- | --- | --- |
| Pregnancies | 57,683 | 8,457 | 49,226 |  |
| Individuals | 38,997 | 7,286 | 34,120 |  |
| Maternal age range, years | 27 – 37 | 29 – 39 | 27 - 37 | <0.001 |
| Race/ethnicity |  |  |  | <0.001 |
| White, n (%) | 34,293 (59%) | 5091 (60%) | 29,202 (59%) | 0.129 |
| Black, n (%) | 6117 (11%) | 1237(15%) | 4880 (10%) | <0.001 |
| Asian, n (%) | 3659 (6%) | 397 (5%) | 3262 (7%) | <0.001 |
| Hispanic ethnicity, n (%) | 7374 (13%) | 951 (11%) | 6423 (13%) | <0.001 |
| Other, n (%) | 6240 (11%) | 781(9%) | 5459 (11%) | <0.001 |
| National ADI score, n | 23 (13, 43) | 23 (13, 43) | 23 (13, 43) | 0.022 |
| State ADI score, n | 4.6 ± 2.1 | 4.6 ± 2.1 | 4.5 ± 2.2 | 0.015 |
| Body mass index, kg/m<sup>2</sup> | 26.2 ± 5.9 | 28.5 ± 7.1 | 25.7 ± 5.6 | <0.001 |
| Systolic blood pressure, mmHg | 113 ± 12 | 119 ± 14 | 112 ± 12 | <0.001 |
| Diastolic blood pressure, mmHg | 70 ± 9 | 74 ± 10 | 69 ± 9 | <0.001 |
| Pulse, bpm | 78 ± 16 | 80 ± 19 | 78 ± 15 | <0.001 |
| Smoking use, n (%) | 2254 (4%) | 405 (5%) | 1849 (4%) | <0.001 |
| Alcohol use, n (%) | 2898 (5%) | 624 (7%) | 2274 (5%) | <0.001 |
| Hemoglobin, g/dL | 12.8 ± 1.0 | 12.8 ± 1.1 | 12.8 ± 1.0 | 0.109 |
| Hypertension medication use, n (%) | 1953 (3%) | 1228 (14%) | 725 (1%) | <0.001 |
| Pre-existing obesity, n (%) | 7068 (12%) | 1684 (20%) | 5384 (11%) | <0.001 |
| Pre-existing diabetes, n (%) | 1786 (3%) | 510 (6%) | 1276 (3%) | <0.001 |
| Pre-existing hypertension, n (%) | 4486 (8%) | 1967 (23%) | 2519 (5%) | <0.001 |
| Pre-existing hyperlipidemia, n (%) | 5953 (10%) | 1099 (13%) | 4854 (10%) | <0.001 |
| Pre-existing CVD, n (%) | 2394 (4%) | 876 (10%) | 1518 (3%) | <0.001 |
| Multimorbidity (>3 CV comorbidities) | 197 (0.3%) | 107 (1.3%) | 90 (0.2%) | <0.001 |
| Prior pregnancy-related cardiovascular complication*, n (%) | 2622 (5%) | 1008 (12%) | 1614 (3%) | <0.001 |
| Gravidity, n | 2.6 ± 1.7 | 2.6 ± 1.8 | 2.6 ± 1.7 | 0.44 |
| Parity, n | 1.3 ± 1.2 | 1.2 ± 1.2 | 1.3 ± 1.2 | 0.12 |
| Estimated gestational age, weeks | 39 ± 4 | 37 ± 4 | 39 ± 3 | <0.001 |

Values are mean ± SD, median (Q1, Q3), or n (%). Values shown exclude missing data. P-value indicates difference between pregnancies with vs without CV complication. Abbreviations: ADI = area deprivation index, PADME = Predictive Analysis with Deep Learning Models for Maternal Endpoints.

### Gestational age, gravidity and parity ascertained via RegEx

Using code-based data alone, estimated gestational age was available for 21,622 (27%) pregnancies. With the application of RegEx, the proportion of pregnancies with available gestational age increased to 74% (**Figure 2**). RegEx more accurately ascertained gestational age than diagnostic codes when compared against manual adjudication (RegEx: r = 0.90, mean absolute error [MAE] = 0.72 weeks, 95% limits of agreements -4.35 to 4.42 weeks; codes: r = 0.83, MAE = 1.68 weeks, 95% limits of agreements -4.73 to 7.93 weeks). Mean estimated gestational age at time of delivery was 39 ± 4 weeks (**Table 1**).

Tabular and code-based data were not available for gravidity and parity. Application of RegEx to clinical notes identified parity and gravidity for 43,637 (76%) of pregnancies. Mean gravidity and parity for pregnancies in PADME was 2.6 ± 1.7 and 1.3 ± 1.2 pregnancies, respectively.

### Prevalence and trends in maternal cardiometabolic comorbidities

Prevalence of maternal cardiometabolic comorbidities in PADME are summarized in **Table 1**. Among 57,683 pregnancies, 12% occurred in the context of pre-existing obesity, 3% with DM, 8% with HTN, and 10% with hyperlipidemia. Mean maternal BMI was 26.2 ± 5.9 kg/m^2^. Prevalence of cardiometabolic comorbidities increased over the study follow-up period (**Figure 3**, **eTable 6**). In age-stratified analyses, prevalence of obesity and hypertension increased over time in all maternal age groups. Hyperlipidemia was stable among pregnancies in younger maternal age groups but increased from 2001-2010 in the older age groups. Diabetes increased across all age groups except in individuals 26-35 years, for whom trends were stable. In age-adjusted analyses, prevalence of maternal obesity increased from 2% in 2001 to 15% in 2019, DM from 1% to 3%, HTN from 3% to 12%, and hyperlipidemia from 3% to 10% (p < 0.01 for all, **eFigure 3**, **eTable 7).**

**Figure 3.**
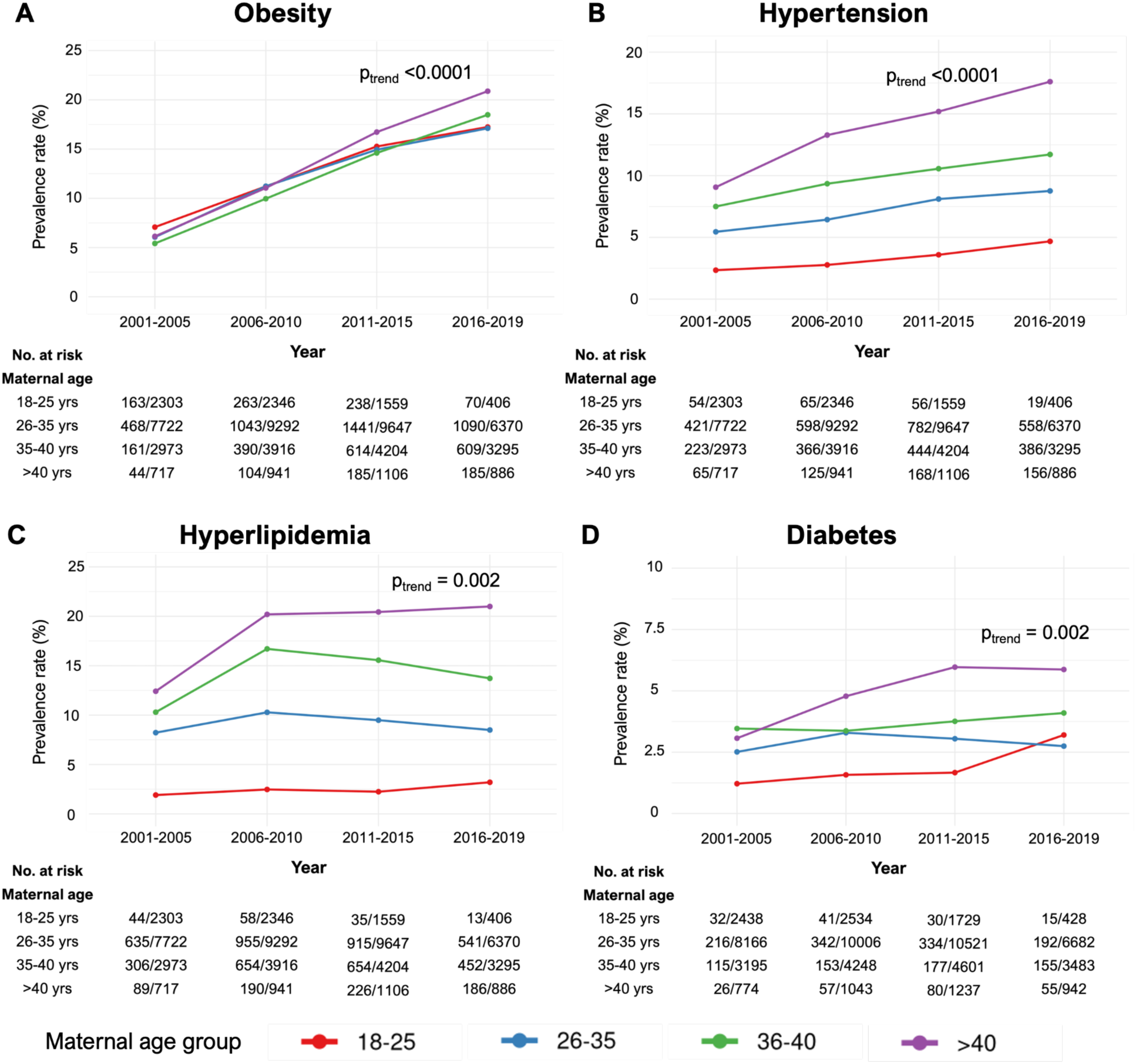
Trends in maternal cardiovascular comorbidities from 2001 to 2019, stratified by maternal age. Displayed are the prevalence rates for preexisting CV comorbidities from 2001 to 2019 in pre-specified time intervals (2001-2005, 2006-2010, 2011-2015, 2016-2019) stratified by maternal age at time of delivery. Cardiovascular comorbidities include obesity (**panel A**), hypertension (**panel B**), diabetes (**panel C**), and hyperlipidemia (**panel D)**.

### Prevalence and trends in existing CVD

Overall prevalence of pre-existing maternal CVD in PADME over the cohort follow-up period was 4% (age-adjusted prevalence: 7%) (**Figure 4**, **eTable 6**). Venous thromboembolism (2%), TIAs (0.5%), and HF (1%) were the most common prevalent maternal CVD conditions. Prevalent arrhythmias including atrial fibrillation, supraventricular tachycardia, and ventricular tachycardia affected 0.7% of pregnancies. Prevalent MI was present in 0.1% of pregnancies.

**Figure 4.**
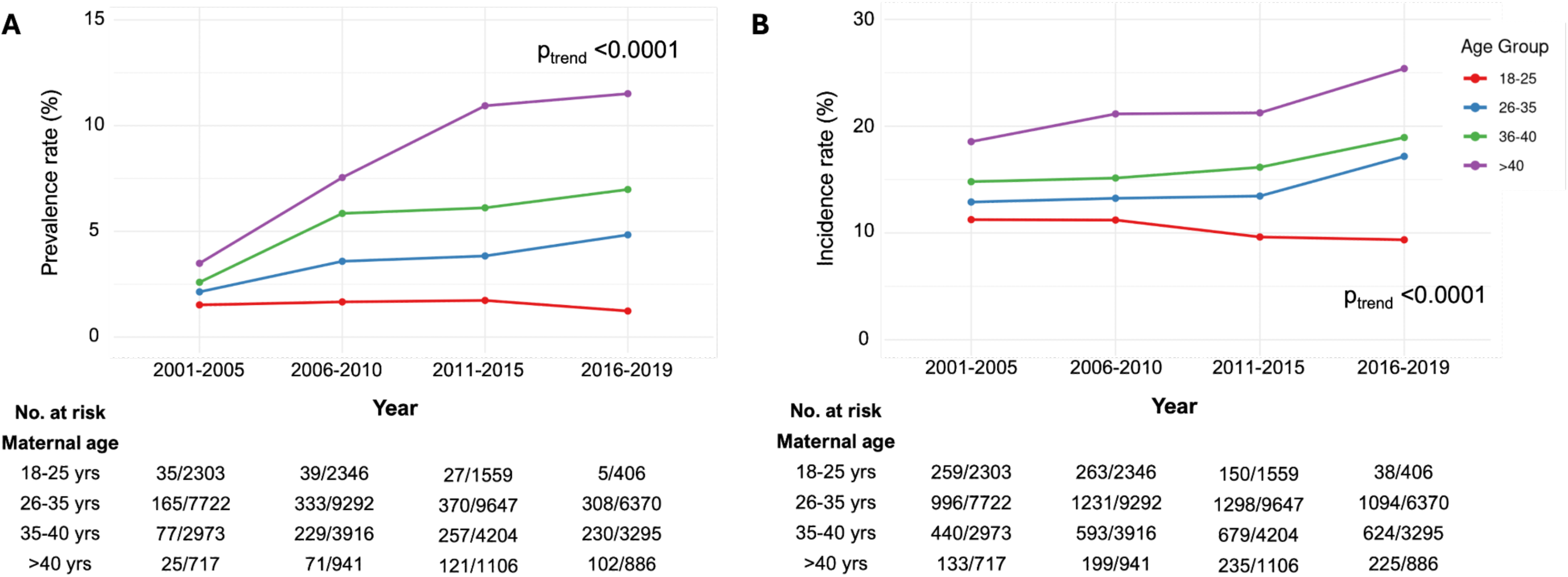
Trends in prevalent and incident pregnancy-related cardiovascular complications from 2001 to 2019. Displayed are the prevalence rates for pre-existing maternal CVD (**panel A**) and incidence rates of pregnancy-related CV complications (**panel B**) from 2001 to 2019 in pre-specified time intervals, stratified by maternal age at time of delivery.

Prevalence of established maternal CVD also increased over the study follow-up period. In age-stratified analyses, prevalence of CVD was lowest among the youngest maternal age group and highest among the oldest (**Figure 4**). Increase over time in prevalent CVD was also greatest among pregnancies in individuals with maternal age >40 years. Overall, age-adjusted prevalence of maternal CVD increased from 1% in 2001 to 7% in 2019 (β 0.05 per 1-year increase, SE 0.004, p-value < 0.0001) (**eFigure 4**, **eTable 7**). Trends were similar in sensitivity analyses restricted to first pregnancies (**eTable 8**).

### Incidence and trends in pregnancy-related cardiovascular complications

Among 57,683 pregnancies, 8,457 (15%) were complicated by an incident CV event including MACE, HDP, or maternal death (**eTable 6**). MACE occurred in 2198 (4%) of pregnancies, with venous thromboembolism as the most common MACE complication during pregnancy (2%) (**eTable 9**). HDP complicated 6678 (12%) of pregnancies. Chronic hypertension occurred in 5% of pregnancies, gestational HTN in 2%, and preeclampsia in 8%. Eclampsia and HELLP syndrome were rare (eclampsia: 0.2%, HELLP syndrome: 0.03%). Maternal deaths were exceedingly rare (n=7, 0.01%) (**eTable 9**).

Incidence of pregnancy-related CV complications increased over the cohort follow-up time (**Figure 4**, **eTable 6**). Age-adjusted incidence of pregnancy-related CV complications increased from 11% in 2001 to 14% in 2019 (β 0.015 per 1-year increase, SE 0.002, p <0.0001, **eFigure 5**, **eTable 7**). Incident HDP also increased from 2001 to 2019 (p <0.001). There was no significant trend in incident MACE over the follow-up period. Results were similar when examining only first pregnancies (**eTable 8**).

### Clinical characteristics and incident pregnancy-related cardiovascular complications

Risk factor profiles differed between pregnancies with and without CV complications. CV complications were more likely to occur among individuals with cardiometabolic risk factors (with vs without CV complication: obesity 20% vs 11%, DM: 6% vs 3%, HTN 23% vs 5%, hyperlipidemia: 13% vs 10%, p-value for all <0.001) and with greater comorbidity burden (with vs without CV complication: >3 CV comorbidities 1.3% vs 0.2%, p-value <0.001, **Table 1**). Incident CV complications during pregnancy were also more likely to occur in individuals with existing CVD (with vs without CV complication: 10% vs 3%, p-value <0.001, **eTable 10**). Finally, among 22,749 PADME multiparous pregnancies, pregnancy-related CV complications were also more frequent in individuals who experienced a pregnancy-related CV complication in ≥ 1 prior pregnancy (with vs without prior pregnancy-related CV complications: 12% vs 3%, p-value <0.001, **Table 1**).

## DISCUSSION

We developed PADME, a multi-institutional EHR-based pregnancy cohort of >57,000 pregnancies with well-curated clinical data and rigorously defined cardiovascular outcomes, and characterized the contemporary epidemiology of pregnancy-related CV complications in a real-world population. Our findings are four-fold. First, the prevalence of maternal cardiometabolic comorbidities and CVD has risen over time. Second, the incidence of cardiovascular complications in pregnancy is appreciable and appears to be increasing. Third, pregnancy-related CV complications were more frequent in the presence of maternal cardiometabolic comorbidities and prevalent CVD. Finally, these findings across a large multi-institutional system were enabled by accurate ascertainment of gravidity, parity, and gestational age from unstructured notes. Taken together, we demonstrate that the contemporary real world burden of maternal cardiometabolic risk factors and subsequent pregnancy-related CV complications are rising in parallel at alarming rates and underscore pregnancy as a critical window of opportunity to optimize CV health.

Our results are consistent with and extend prior studies that show that CV health of individuals entering pregnancy has been declining over the past several decades.^19–21^ Previous studies have estimated that preexisting CVD affects 1-4% of all pregnancies and that the prevalence has risen in recent decades.^21,22^ In a retrospective study of hospitalized pregnant patients, the age-adjusted prevalence of CVD was 11.3% and increased from 9.2% in 2010 to 14.8% in 2019.^19^ We show similar prevalence estimates of maternal CVD (crude prevalence 4%, age-adjusted prevalence 7%) and rising prevalence rates of both maternal cardiometabolic comorbidities and CVD over the first two decades of the 21st century.^23^

We now demonstrate that the incidence of CV complications during pregnancy is also substantial and increasing, even among individuals without a history of CVD. Few studies have comprehensively examined incidence of pregnancy-related CV complications beyond peripartum cardiomyopathy and HDP, and the available studies were conducted in small samples restricted to individuals with existing CVD.^12,22,24^ For example, the Registry of Pregnancy and Cardiac Disease (ROPAC), one of the largest pregnancy cohorts with prospectively ascertained cardiovascular outcome data, included only 1321 pregnancies, of which 57% had congenital heart disease.^10^ The highly selective nature of these cohorts has precluded the understanding of the population-level burden of pregnancy-related complications, particularly among individuals without established CVD. Our study contributes robust estimates of rising incidence of pregnancy-related CV complications from >57,000 pregnancies, 96% without known CVD, a notable strength as most adverse CV events in pregnancy occur in the context of previously undiagnosed CVD.

We further demonstrated that CV complications were more common in pregnant individuals with CV comorbidities and/or history of CVD, highlighting pre-pregnancy CV health optimization as a key provision needed to improve maternal outcomes. Early studies of CVD in pregnancy showed that maternal CVD is a significant risk factor for maternal morbidity and mortality, and risk stratified specific CV lesions to help guide preconception counseling.^8,10^ We and others now show that maternal cardiometabolic comorbidities, even in the absence of existing CVD, confer significant risk of pregnancy-related CV complications.^25^ In PADME, individuals with multimorbidity had nearly 7x higher incidence of pregnancy-related CV complications compared with individuals with ≤ 3 maternal CV comorbidities.

Finally, the design of PADME represents an important methodological advance. Large datasets like EHRs or insurance claims databases have been underutilized for pregnancy outcomes due to challenges related to ascertainment bias, data missingness, and accurate identification of pregnancy episodes.^26–28^ To address these limitations, we first identified pregnancies from a cohort of individuals receiving regular primary care to reduce ascertainment bias and enable longitudinal follow-up.^15^ Second, we employed machine learning approaches to recover missing data, including vital signs and specific pregnancy-related features like gestational age that are not routinely captured by diagnostic codes or tabular data.^29,30^ In PADME, RegEx recovered the gestational age for 74% of pregnancy encounters with superior accuracy to diagnostic codes, increasing our cohort size from N=21,622 to N=57,683 pregnancies, 20-50x the scale of existing pregnancy cohorts. Finally, to ensure rigorous outcome ascertainment in PADME, we utilized and refined validated algorithms to define presence of disease and confirmed accuracy of our selection methods via manual adjudication.^15,31^

### Limitations

Our study has several limitations. First, we identified pregnancy encounters using EHR-based pregnancy codes. Although manual validation of candidate pregnancy encounters demonstrated excellent accuracy and agreement, we acknowledge that code-based algorithms may be imperfectly sensitive. Second, analyses of unstructured text were used to minimize missingness of key clinical variables such as vital sign data, but missingness of other covariates that proved more difficult to extract (e.g. alcohol use, smoking status) remains considerable. Third, pregnancy encounters identified in the MGB system occurred in tertiary or quaternary care settings, while many deliveries occur in community practice or outpatient settings. Finally, while PADME includes larger absolute numbers of individuals of color compared with other contemporary cohorts, 59% of individuals are White. Generalizability to populations with greater racial/ethnic diversity is an important focus of future work.

## CONCLUSIONS

In a novel contemporary EHR-based pregnancy cohort comprising over 57,000 pregnancies within a large multi-institutional healthcare system, we found that CV complications occurred in 15% of pregnancies among individuals with and without established CVD. The burden of maternal cardiometabolic comorbidities has increased substantially over the past 2 decades, along with a concomitant rise in incident pregnancy-related CV complications. Together, our findings showcase an alarming trend of rising real-world burden of pregnancy-related CV complications and highlights pregnancy from preconception to the postpartum period as a critical window of opportunity to implement primary prevention strategies and optimize CV health.

## Data Availability

All data produced in the present work are contained in the manuscript

## ABBREVIATIONS

CPT: Current Procedural Terminology
CV: cardiovascular
CVD: cardiovascular disease
EHR: electronic health record
HDP: hypertensive disorders of pregnancy
HELLP: hemolysis, elevated liver enzymes, low platelets
HF: heart failure
ICD: International Classification of Disease
MGB: Mass General Brigham
MI: myocardial infarction
NLP: natural language processing
PADME: Predictive Analysis with Deep Learning Models for Maternal Endpoints
RegEx: regular expression
TIA: transient ischemic attack
US: United States

## REFERENCES

1. Howell EA. Reducing Disparities in Severe Maternal Morbidity and Mortality. Clin Obstet Gynecol. 2018;61(2):387. doi:10.1097/GRF.0000000000000349

2. Fleszar LG, Bryant AS, Johnson CO, et al. Trends in State-Level Maternal Mortality by Racial and Ethnic Group in the United States. JAMA. 2023;330(1):52–61. doi:10.1001/jama.2023.9043

3. Maternal Mortality Rates in the United States, 2021. March 16, 2023. Accessed July 22, 2024. https://www.cdc.gov/nchs/data/hestat/maternal-mortality/2021/maternal-mortality-rates-2021.htm

4. Maternal Mortality in the United States: A Primer. doi:10.26099/ta1q-mw24

5. Mehta LS, Sharma G, Creanga AA, et al. Call to Action: Maternal Health and Saving Mothers: A Policy Statement From the American Heart Association. Circulation. 2021;144(15):e251–e269. doi:10.1161/CIR.0000000000001000

6. CDC. Pregnancy Mortality Surveillance System. Maternal Mortality Prevention. May 20, 2024. Accessed October 28, 2024. https://www.cdc.gov/maternal-mortality/php/pregnancy-mortality-surveillance/index.html

7. Creanga AA, Syverson C, Seed K, Callaghan WM. Pregnancy-Related Mortality in the United States, –2013. Obstet Gynecol. 2017;130(2):366. doi:10.1097/AOG.0000000000002114

8. Silversides CK, Grewal J, Mason J, et al. Pregnancy Outcomes in Women With Heart Disease: The CARPREG II Study. J Am Coll Cardiol. 2018;71(21):2419–2430. doi:10.1016/j.jacc.2018.02.076

9. Siu SC, Sermer M, Colman JM, et al. Prospective multicenter study of pregnancy outcomes in women with heart disease. Circulation. 2001;104(5):515–521. doi:10.1161/hc3001.093437

10. Roos-Hesselink J, Baris L, Johnson M, et al. Pregnancy outcomes in women with cardiovascular disease: evolving trends over 10 years in the ESC Registry Of Pregnancy And Cardiac disease (ROPAC). Eur Heart J. 2019;40(47):3848–3855. doi:10.1093/eurheartj/ehz136

11. Pfaller B, Sathananthan G, Grewal J, et al. Preventing Complications in Pregnant Women With Cardiac Disease. J Am Coll Cardiol. 2020;75(12):1443–1452. doi:10.1016/j.jacc.2020.01.039

12. van Hagen IM, Boersma E, Johnson MR, et al. Global cardiac risk assessment in the Registry Of Pregnancy And Cardiac disease: results of a registry from the European Society of Cardiology. Eur J Heart Fail. 2016;18(5):523–533. doi:10.1002/ejhf.501

13. Hagen IM van, Roos-Hesselink JW. Pregnancy in congenital heart disease: risk prediction and counselling. Heart. 2020;106(23):1853–1861. doi:10.1136/heartjnl-2019-314702

14. MacDonald SC, Cohen JM, Panchaud A, McElrath TF, Huybrechts KF, Hernández-Díaz S. Identifying Pregnancies in Insurance Claims Data: Methods and Application to Retinoid Teratogenic Surveillance. Pharmacoepidemiol Drug Saf. 2019;28(9):1211–1221. doi:10.1002/pds.4794

15. Khurshid S, Reeder C, Harrington LX, et al. Cohort design and natural language processing to reduce bias in electronic health records research. NPJ Digit Med. 2022;5(1):47. doi:10.1038/s41746-022-00590-0

16. CDC. Pregnancy-Related Deaths: Data From Maternal Mortality Review Committees in 36 U.S. States, 2017–2019. Maternal Mortality Prevention. May 30, 2024. Accessed November 8, 2024. https://www.cdc.gov/maternal-mortality/php/data-research/mmrc-2017-2019.html

17. Curtin LR, Klein RJ. Direct Standardization (Age-Adjusted Death Rates): (584012012-001). Published online 1995. doi:10.1037/e584012012-001

18. Welcome to Python.org. Python.org. July 18, 2024. Accessed July 22, 2024. https://www.python.org/

19. Majmundar M, Doshi R, Patel KN, Zala H, Kumar A, Kalra A. Prevalence, trends, and outcomes of cardiovascular diseases in pregnant patients in the USA: 2010-19. Eur Heart J. 2023;44(9):726–737. doi:10.1093/eurheartj/ehac669

20. Elkayam U, Goland S, Pieper PG, Silverside CK. High-Risk Cardiac Disease in Pregnancy: Part I. J Am Coll Cardiol. 2016;68(4):396–410. doi:10.1016/j.jacc.2016.05.048

21. Sliwa K, Böhm M. Incidence and prevalence of pregnancy-related heart disease. Cardiovasc Res. 2014;101(4):554–560. doi:10.1093/cvr/cvu012

22. Bruno AM, Allshouse AA, Metz TD, Theilen LH. Trends in Hypertensive Disorders of Pregnancy in the United States From 1989 to 2020. Obstet Gynecol. 2022;140(1):83–86. doi:10.1097/AOG.0000000000004824

23. Ogden CL, Carroll MD, Fryar CD, Flegal KM. Prevalence of Obesity Among Adults and Youth: United States, 2011-2014. NCHS Data Brief. 2015;(219):1–8.

24. Kuklina EV, Callaghan WM. Cardiomyopathy and other myocardial disorders among hospitalizations for pregnancy in the United States: 2004-2006. Obstet Gynecol. 2010;115(1):93–100. doi:10.1097/AOG.0b013e3181c4ee8c

25. Zahid S, Jha S, Kaur G, et al. Parccs. JACC Adv. 2024;3(8):101095. doi:10.1016/j.jacadv.2024.101095

26. Cowie MR, Blomster JI, Curtis LH, et al. Electronic health records to facilitate clinical research. Clin Res Cardiol Off J Ger Card Soc. 2017;106(1):1–9. doi:10.1007/s00392-016-1025-6

27. Kaplan RM, Chambers DA, Glasgow RE. Big data and large sample size: a cautionary note on the potential for bias. Clin Transl Sci. 2014;7(4):342–346. doi:10.1111/cts.12178

28. Haneuse S, Daniels M. A General Framework for Considering Selection Bias in EHR-Based Studies: What Data Are Observed and Why? EGEMS Wash DC. 2016;4(1):1203. doi:10.13063/2327-9214.1203

29. Margulis AV, Setoguchi S, Mittleman MA, Glynn RJ, Dormuth CR, Hernández-Díaz S. Algorithms to estimate the beginning of pregnancy in administrative databases. Pharmacoepidemiol Drug Saf. 2013;22(1):16–24. doi:10.1002/pds.3284

30. Sarayani A, Wang X, Thai TN, Albogami Y, Jeon N, Winterstein AG. Impact of the Transition from ICD-9-CM to ICD-10-CM on the Identification of Pregnancy Episodes in US Health Insurance Claims Data. Clin Epidemiol. 2020;12:1129–1138. doi:10.2147/CLEP.S269400

31. Khurshid S, Keaney J, Ellinor PT, Lubitz SA. A Simple and Portable Algorithm for Identifying Atrial Fibrillation in the Electronic Medical Record. Am J Cardiol. 2016;117(2):221–225. doi:10.1016/j.amjcard.2015.10.031

