## Supplemental Appendix for "Contemporary Burden of Cardiovascular Disease in Pregnancy: Insights from a Real-World Pregnancy Electronic Health Record Cohort"

**Supplementary Material**

### eMethods. Supplemental Methods

#### Data ingestion pipeline

We obtained a comprehensive range of EHR data from 1 year prior to start of pregnancy to 1 year postpartum including demographics, vital signs and anthropometrics, narrative notes, laboratory results, medication lists, and radiology/cardiology diagnostic test reports for a preliminary set of 111,143 pregnancies (54,289 individuals) identified for inclusion in the PADME cohort through the Research Patient Data Registry (RPDR, Boston, MA), a data repository of complete EHR data for all individuals receiving care within MGB.[17] Data elements were then processed using the standardized data ingestion pipeline (the "JEDI" Extractive Data Infrastructure) which integrates distinct files containing an array of EHR data types into a unified, indexed file system (Hierarchical Data Format 5) as previously described.[15,18]

#### Ascertaining gravidity, parity, and gestational age from unstructured clinical notes using regular expressions

To extract gravidity, parity, and gestational age from unstructured clinical notes, we utilized regular expressions (RegEx) tailored to identify specific patterns commonly found in clinical documentation. The process involved the following steps:

*Pattern Identification and Extraction:* To identify relevant patterns for extracting gestational age, we analyzed a sample of unstructured clinical notes to understand how information is typically documented. Patterns included variants such as gravida 4 para 2 and G4P2012 for gravidity and parity, and 34wn7d, 34 weeks and 3 days, and 34 3/7 weeks for gestational age.

*Iterative Refinement:* We refined the RegEx through the following process:

- Initial Testing: The patterns were applied to a subset of clinical notes to test their performance in capturing the desired information.
- Error Analysis: We manually reviewed mismatches or missed extractions to identify cases where the initial expressions failed (e.g., variations in spacing, alternative wording, or inconsistent formats).

- RegEx Adjustments: The expressions were iteratively adjusted to improve their robustness, such as accommodating alternate delimiters (w vs. wk), spaces between components, or fractional weeks written in different formats.
- Adding Flexibility: Additional patterns were included to cover edge cases identified during testing (e.g., "34w7d" without the "n").

We employed the following RegEx terms to ascertain gravidity and parity:

- For extracting patterns like “gravida 4, para 2”: r'(?P<gravida>Gravida\s*[0-9]+)\s*(?P<text>.*?)\s*(?P<para>Para\s*[0-9]+)'
- For extracting patterns like “G4P2012/ G4P2”: r'(?P<gpg>G\d+(?:\s*)P\d+)'

We employed the following RegEx terms to ascertain gestational age:

- For extracting patterns like 34wn7d: r'(?P<nwnd>[0-9]+w[0-9]+d)'
- For extracting patterns like 34 3/7 weeks: r'(?P<weeks_frac>[0-9]+ and [0-9]+/7 weeks)'
- For extracting patterns like 34+3 weeks gestation: r'(?P<at_gestation_plus_num>[0-9]+\s*\+\s*[0-9])(?P<at_gestation_plus> weeks gestation)'
- For extracting patterns like 34 +3/7 weeks: r'(?P<weeks_plus_frac>[0-9]+\s*[+]?\s*[0-9]+/7 weeks)'
- For extracting patterns like Intrauterine pregnancy at x x/7 weeks: r'(?P<iup_frac>Intrauterine\s*(?:uterine)\s*pregnancyat\s*)(?P<iup_frac_age>[0-9]+\s*[-]?\s*[0-9]+\/[0-9]\s*weeks)'
- r'(?P<gestation>Gestational age at time of delivery:\s*)(?P<gestation_age>[0-9]+[.]?[0-9]*)'
- r'(?P<gestation_v>Gestational age at visit:\s*)(?P<gestationv_age>[0-9]+[.]?[0-9]*)'
- r'(?P<at_gestation_num>[0-9]+)(?P<at_gestation> weeks gestation)'

**Ascertaining Gestational Age**

We employed the following stepwise approach to estimate the pregnancy episode (**eFigure 1**). The first step identifies the date of the pregnancy endpoint ($T_1$), defined as the date associated with a given pregnancy endpoint code. For pregnancy encounters with multiple pregnancy delivery codes, the date of the first observed code was considered the pregnancy delivery date. To prevent misclassification of consecutive pregnancy delivery codes assigned to a single pregnancy episode as distinct pregnancy episodes, we also applied a relevant washout period between consecutive pregnancy endpoint codes as follows: 200 days for live births, unclassified deliveries, and stillbirth and 60 days for spontaneous abortion, induced abortions, and ectopic pregnancies.

The second step identifies the date of a clinical note with documented gestational age ($T_{note}$). This third step identifies the gestational age at the time of the pregnancy endpoint ($G_1$). For instances where the clinical note with documented gestational age occurs before the pregnancy endpoint ($T_{note} < T_1$), the gestational age at time of pregnancy endpoint is calculated by adding the gestational age documented in the clinical note to the time period between pregnancy endpoint and the clinical note ($G_1 = G_{note} + [T_{note} - T_1]$). For instances where the clinical note with documented gestational age occurs after the pregnancy ($T_{note} > T_1$), the gestational age documented in the clinical note is assigned as the gestational age at the time of pregnancy endpoint ($G_1 = G_{note}$). When there are multiple notes with documented gestational age, the final gestational age of the pregnancy endpoint will be anchored to the clinical note that is most proximal in time to the pregnancy endpoint. The final step defines the start of the pregnancy period which is calculated by subtracting the gestational age at the time of the pregnancy endpoint from the date of the pregnancy endpoint ($T_0 = T_1 - G_1$).

**Defining pregnancy episodes**

For pregnancy encounters with multiple pregnancy endpoint codes, the date of the first observed code was considered the delivery date. To prevent misclassification of consecutive pregnancy delivery codes assigned to a single pregnancy episode as distinct pregnancy episodes, we applied a 200 day blanking period between consecutive pregnancy delivery codes. The date of the estimated last menstrual period, or gestational age 0, was defined as the pregnancy delivery date minus the gestational age at time of delivery.

**Assessment of baseline clinical variables**

- *Demographic data:* Baseline demographic data including age, sex, race, and zip code were obtained from dedicated demographic fields from the EHR.
- *Vital signs:* Vital signs including weight, height, body mass index (kg/m$^2$), systolic and diastolic blood pressures, and pulse were extracted from clinical encounters. Due to significant missingness (>40%) of

baseline vital signs in the EHR, a NLP algorithm was used to recover vital signs from unstructured notes as previously described.[15]

- *Prevalent comorbidities and disease*: Comorbidities and disease conditions of interest were selected based on established or suspected associations with CV complications during pregnancy. Prevalent comorbidities of interest included obesity, diabetes (DM), hypertension (HTN), and hyperlipidemia. Prevalent disease conditions of interest included myocardial infarction (MI), heart failure (HF), vascular dissection, thromboembolism (venous, pulmonary and systemic), cerebrovascular disease (including transient ischemic attacks [TIA], hemorrhagic cerebrovascular accidents [CVA], and ischemic CVA), ventricular and atrial arrhythmias, and cardiac arrest. Disease-related covariates were ascertained based on presence of at least one ICD, CPT, and/or EHR-specific diagnosis code corresponding to the relevant disease (**eTable 2**).[15,19]
- *Medication use*: HTN medication use was ascertained using medication lists (**eTable 3**).
- *Smoking status*: Smoking status was ascertained from formal smoking questionnaire data documented in the EHR and was stratified into past or present smoker or never smoker. The assessment period spanned any time period prior to pregnancy endpoint. Smoking status was marked as missing if data on smoking status were not available during the assessment period.
- *Alcohol use*: Alcohol use was obtained from formal alcohol questionnaire data in the EHR and was defined as yes or no. The assessment period was restricted to the pregnancy episode (from estimated gestational age 0 to the pregnancy endpoint). Alcohol use was marked as missing if alcohol data were not available during the assessment period.
- *Laboratory data*: Laboratory values including basic metabolic panel, complete blood count, liver function tests, and thyroid function tests were obtained from dedicated laboratory fields from the EHR.
- *Socioeconomic status:* Socioeconomic status was captured using the Area Deprivation Index (ADI), a validated surrogate marker of national and state level socioeconomic disadvantage based on an individual patient's zip code.[20,21] The ADI is a composite measure created by the Health Resources and Services Administration that integrates 17 measures of employment, income, housing, and education identified from the American Community Survey. As the MGB EHR only captures 5-digit zip code patient data, we used averaged ADI scores across all 9-digit zip codes for a given 5-digit zip code area.

**eTable 1.** Pregnancy endpoint definitions

| **Category** | **ICD-9, ICD-10, and CPT codes** | **Code Description** |
|---|---|---|
| Live birth | V30.xx, V31.xx, V32.xx, V34.xx, V35.xx, V36.xx, V37.xx, V39.xx, V27.0, V27.2, Z38.xx, Z35.5x, Z37.2, Z37.0, O60.23xx, O60.22xx, O75.82 | Single liveborn, Twin birth, Twin birth - mate stillborn, Other multiple birth, Other multiple birth - all mates stillborn, Other multiple birth - mates stillborn and liveborn, Other multiple birth - unspecified whether mates stillborn or liveborn, Liveborn - unspecified whether single, twin or multiple, Mother with single liveborn, Mother with twins both liveborn, Multiple liveborn infant, Multiple births - all liveborn, Twins - both liveborn, Single live birth, Term delivery with preterm labor - third trimester, Term delivery with preterm labor - second trimester, Onset (spontaneous) of labor after 37 completed weeks of gestation but before 39 completed weeks gestation - with delivery by (planned) cesarean section |
| Mixed birth | V27.3, V27.6, Z37.6x | Mother with twins - one liveborn and one stillborn, Mother with other multiple births - some liveborn, Multiple births - some liveborn, |
| Ectopic pregnancy | 633.xx, O00.x, 74.3, 59120, 59121, 59130, 59135, 59136, 59140, 59150, 59151 | Ectopic pregnancy, Ectopic pregnancy, Removal Of Extra Tubal Ectopic Pregnancy, Surgical treatment of ectopic pregnancy; tubal or ovarian - requiring salpingectomy and/or oophorectomy, Surgical treatment of ectopic pregnancy; tubal or ovarian - without salpingectomy and/or oophorectomy, Surgical treatment of ectopic pregnancy; abdominal pregnancy, Surgical treatment of ectopic pregnancy; interstitial uterine pregnancy requiring total hysterectomy, Surgical treatment of ectopic pregnancy; interstitial uterine pregnancy with partial resection of uterus, Surgical treatment of ectopic pregnancy; cervical with evacuation, Laparoscopic treatment of ectopic pregnancy; without salpingectomy and/or oophorectomy, Laparoscopic treatment of ectopic pregnancy; with salpingectomy and/or oophorectomy |
| Stillbirth | 88016, S226x, V27.1, V27.4, V27.7, 656.4x, O36.4xxx, Z37.1, Z37.4, Z37.7 | Necropsy (autopsy) - gross examination only; macerated stillborn, Induced abortion - 25 weeks or greater, Mother with single stillborn, Mother with twins both stillborn, Mother with other multiple birth all stillborn, Intrauterine death affecting management of mother, Maternal care for intrauterine death, Single stillbirth, Twins - both stillborn, Other multiple births - all stillborn |
| Spontaneous abortion | 634.xx, 632, 631, 637.xx, 1956, 59812, 59820, 59821, 59830, O02.1, O02.81, | Spontaneous abortion, Missed abortion, Inappropriate change in quantitative human chorionic gonadotropin (hCG) in early pregnancy, Unspecified abortion |

| | | |
|---|---|---|
| | O02.0, O02.89, O02.9, O03.6, O03.1, O03.8x, O03.3x, O03.7, O03.2, O03.9, O03.4 | complicated by genital tract and pelvic infection, Anesthesia for incomplete or missed abortion procedures, Treatment of incomplete abortion - any trimester - completed surgically, Treatment of missed abortion - completed surgically; first trimester, Treatment of missed abortion - completed surgically; second trimester, Treatment of septic abortion - completed surgically, Missed abortion, Inappropriate change in quantitative human chorionic gonadotropin (hCG) in early pregnancy, Blighted ovum and non hydatidiform mole, Other abnormal products of conception, Abnormal product of conception - unspecified, Delayed or excessive hemorrhage following complete or unspecified spontaneous abortion, Delayed or excessive hemorrhage following incomplete spontaneous abortion, Damage to organs following spontaneous abortion, Renal failure following spontaneous abortion, Embolism following complete or unspecified spontaneous abortion, Embolism following incomplete spontaneous abortion, Incomplete spontaneous abortion without complication |
| Induced abortion | 1966, 59840, 59841, 5985x, S2260, 635.xx, 636.xx, 779.6, 75, 69.01, 69.51, 74.91, O04.8x, O04.7, O04.6, O04.5, Z33.2, 10A0xZZ | Anesthesia for induced abortion procedures, Induced abortion - by dilation and curettage, Induced abortion - by dilation and evacuation, Induced abortion - by 1 or more intra-amniotic injections (amniocentesis-injections); including hospital admission and visits, delivery of fetus and secundines, Induced abortion - 17 to 24 weeks, Legally induced abortion, Illegal abortion, Termination of pregnancy (fetus), Intra-Amniotic Injection For Abortion, Dilation And Curettage For Termination Of Pregnancy, Aspiration Curettage Of Uterus For Termination Of Pregnancy, Hysterotomy To Terminate Pregnancy, Damage to organs following (induced) termination of pregnancy, Embolism following (induced) termination of pregnancy, Genital tract and pelvic infection following (induced) termination of pregnancy, Delayed or excessive hemorrhage following (induced) termination of pregnancy, Encounter for elective termination of pregnancy, Abortion of Products of Conception |
| Unclassified delivery | V33.xx, 641.xx, 643.0x-643.1x, 643.2x-643.9x, 644.21, 645.11, 645.21, 646.0x - 646.1x, 646.22, 646.31, 646.4x, 646.5x, 646.6x, 646.71, 646.8x, 646.91, 647.xx, 648.xx-649.xx, 650, 651.xx, 652.xx, 653.xx, 654.xx, 655.xx, 656.xx, 658.xx, 659.xx, 660.xx, 661.xx, 662.xx, 663.xx, 664.xx, 665.xx, 667.xx, 668.xx, 669.xx, 670.xx, 671.xx, 673.xx, | Twin birth - unspecified whether mate liveborn or stillborn, Placenta previa - delivered with or without mention of antepartum condition, Hyperemesis gravidarum - delivered with or without mention of antepartum condition, Vomiting complications in pregnancy - delivered with or without mention of antepartum condition, Early onset of delivery - delivered with or without mention of antepartum condition, Post term pregnancy - delivered with or without mention of antepartum condition, Prolonged pregnancy - delivered with or without mention of antepartum condition, Weight issues - delivered with or without mention of antepartum condition, Unspecified renal disease in pregnancy - without mention of hypertension; delivered with mention of postpartum complication, Recurrent |

| | | |
|---|---|---|
| | 674.xx, 675.xx- 676.xx, 678.01, 678.11, 679.xx, O82, O99.xxx, O77.x, O80, O70.x, O74.xx-O75.xx, O69.9xxx, O99.8xx, O88.xx, 72.x, 73.x, 74.x, 196x, 59409, 59514, 59612, 59620 | pregnancy loss - delivered with or without mention of antepartum condition, Peripheral neuritis in pregnancy - delivered with or without mention of antepartum/postpartum condition, Asymptomatic bacteriuria in pregnancy - delivered with or without mention of antepartum/postpartum condition, Infections of genitourinary tract in pregnancy - delivered with or without mention of antepartum/postpartum condition, Liver and biliary tract disorders in pregnancy - delivered with or without mention of antepartum condition, Other specified complications of pregnancy - delivered with or without mention of antepartum condition, Unspecified complication of pregnancy - delivered with or without mention of antepartum condition, Diseases of mother complicating pregnancy and childbirth - delivered with mention of postpartum complication, Other current conditions of mother complicating pregnancy and childbirth - delivered, with or without mention of antepartum condition, Normal delivery, Multiple pregnancies - delivered with or without mention of antepartum condition, Unstable presentation - delivered with or without mention of antepartum condition, Contraction of pelvis - delivered with or without mention of antepartum condition, Abnormalities of uterus - delivered with or without mention of antepartum condition, Abnormality in fetus-affecting management of mother - delivered with or without mention of antepartum condition, Fetal distress - affecting management of mother - delivered with or without mention of antepartum condition, Delayed delivery after spontaneous or unspecified rupture of membranes - delivered with or without mention of antepartum condition, Failed induction of labor - delivered - with or without mention of antepartum condition, Obstruction by during labor - delivered with or without mention of antepartum condition, Primary uterine inertia - delivered with or without mention of antepartum condition, Prolonged labor - delivered with or without mention of antepartum condition, Cord complicating labor and delivery - delivered with or without mention of antepartum condition, Perineal laceration - delivered with or without mention of antepartum condition, Rupture of uterus during labor - delivered with or without mention of antepartum condition, Retained placenta without hemorrhage - delivered with mention of postpartum complication, Pulmonary complications of anesthesia or other sedation in labor and delivery - delivered with or without mention of antepartum condition, Maternal distress complicating labor and delivery - delivered with or without mention of antepartum condition, Puerperal complications - delivered with mention of postpartum complication, Venous complications of pregnancy and the puerperium - delivered with or without mention of antepartum condition, Embolism complications - delivered with mention of postpartum complication, Cerebrovascular disorders in the puerperium - delivered with mention of postpartum complication, Complications associated with breast affecting childbirth - delivered with or |

| | | |
|---|---|---|
| | | without mention of antepartum condition, Fetal hematologic conditions - delivered with or without mention of antepartum condition, Fetal conjoined twins - delivered with or without mention of antepartum condition, Maternal complications from in utero procedure - delivered with or without mention of antepartum condition, Encounter for cesarean delivery without indication, Conditions complicating childbirth, Labor and delivery complicated by fetal stress - unspecified, Encounter for full-term uncomplicated delivery, Perineal laceration during delivery - unspecified, Other specified complications of labor and delivery, Labor and delivery complicated by cord complication, Other infection carrier state complicating childbirth, Other embolism in childbirth, Forceps Operation/ Vacuum Extraction, Other Operations Assisting Delivery, Classical Cesarean Section, Anesthesia for vaginal delivery only, Vaginal delivery only (with or without episiotomy and/or forceps, Cesarean delivery only, Vaginal delivery only, after previous cesarean delivery (with or without episiotomy and/or forceps); including postpartum care, Cesarean delivery only, following attempted vaginal delivery after previous cesarean delivery; including postpartum care |

**eTable 2.** Comorbidity and disease condition definitions

| Phenotype | Code Type | Data codes | Data code definitions |
|---|---|---|---|
| Diabetes | ICD-9, ICD-10 | 249, 249.01, 249.1, 249.11, 249.2, 249.21, 249.3, 249.31, 249.4, 249.41, 249.5, 249.51, 249.6, 249.61, 249.7, 249.71, 249.8, 249.81, 249.9, 249.91, 250, 250.01, 250.02, 250.03, 250.1, 250.11, 250.12, 250.13, 250.2, 250.21, 250.22, 250.23, 250.3, 250.31, 250.32, 250.33, 250.4, 250.41, 250.42, 250.43, 250.5, 250.51, 250.52, 250.53, 250.6, 250.61, 250.62, 250.63, 250.7, 250.71, 250.72, 250.73, 250.8, 250.81, 250.82, 250.83, 250.9, 250.91, 250.92, 250.93, 357.2, 362.01, 362.02, 362.03, 362.04, 362.05, | Secondary diabetes mellitus without mention of complication, not stated as uncontrolled, or unspecified, Secondary diabetes mellitus without mention of complication, uncontrolled, Secondary diabetes mellitus with ketoacidosis, not stated as uncontrolled, or unspecified, Secondary diabetes mellitus with ketoacidosis, uncontrolled, Secondary diabetes mellitus with hyperosmolarity, not stated as uncontrolled, or unspecified, Secondary diabetes mellitus with hyperosmolarity, uncontrolled, Secondary diabetes mellitus with other coma, not stated as uncontrolled, or unspecified, Secondary diabetes mellitus with other coma, uncontrolled, Secondary diabetes mellitus with renal manifestations, not stated as uncontrolled, or unspecified, Secondary diabetes mellitus with renal manifestations, uncontrolled, Secondary diabetes mellitus with ophthalmic manifestations, not stated as uncontrolled, or unspecified, Secondary diabetes mellitus with ophthalmic manifestations, uncontrolled, Secondary diabetes mellitus with neurological manifestations, not stated as uncontrolled, or unspecified, Secondary diabetes mellitus with neurological manifestations, uncontrolled, Secondary diabetes mellitus with peripheral circulatory disorders, not stated as uncontrolled, or unspecified, Secondary diabetes mellitus with peripheral circulatory disorders, uncontrolled, Secondary diabetes mellitus with other specified manifestations, not stated as uncontrolled, or unspecified, Secondary diabetes mellitus with other specified manifestations, uncontrolled, Secondary diabetes mellitus with unspecified complication, not stated as uncontrolled, or unspecified, Secondary diabetes mellitus with unspecified complication, uncontrolled, Diabetes mellitus without mention of complication, type II or unspecified type, not stated as uncontrolled, Diabetes mellitus without mention of complication, type I [juvenile type], not stated as uncontrolled, Diabetes mellitus without mention of complication, type II or unspecified type, uncontrolled, Diabetes mellitus without mention of complication, type I [juvenile type], uncontrolled, Diabetes with ketoacidosis, type II or unspecified type, not stated as uncontrolled, Diabetes with ketoacidosis, type I [juvenile type], not stated as uncontrolled, Diabetes with ketoacidosis, type II or unspecified type, uncontrolled, Diabetes with ketoacidosis, type I [juvenile type], uncontrolled, Diabetes with hyperosmolarity, type II or unspecified type, not stated as uncontrolled, Diabetes with hyperosmolarity, type I [juvenile type], not stated as uncontrolled, Diabetes with hyperosmolarity, type II or unspecified type, uncontrolled, Diabetes with hyperosmolarity, type I [juvenile type], uncontrolled, Diabetes with other coma, type II |

| | | | |
|---|---|---|---|
| | | 362.06, 362.07, 366.41, 791.6, E08.00, E08.01, E08.10, E08.11, E08.21, E08.22, E08.29, E08.311, E08.319, E08.321, E08.329, E08.331, E08.339, E08.341, E08.349, E08.351, E08.359, E08.36, E08.39, E08.40, E08.41, E08.42, E08.43, E08.44, E08.49, E08.51, E08.52, E08.610, E08.618, E08.620, E08.621, E08.622, E08.628, E08.630, E08.638, E08.641, E08.649, | or unspecified type, not stated as uncontrolled, Diabetes with other coma, type I [juvenile type], not stated as uncontrolled, Diabetes with other coma, type II or unspecified type, uncontrolled, Diabetes with other coma, type I [juvenile type], uncontrolled, Diabetes with renal manifestations, type II or unspecified type, not stated as uncontrolled, Diabetes with renal manifestations, type I [juvenile type], not stated as uncontrolled, Diabetes with renal manifestations, type II or unspecified type, uncontrolled, Diabetes with renal manifestations, type I [juvenile type], uncontrolled, Diabetes with ophthalmic manifestations, type II or unspecified type, not stated as uncontrolled, Diabetes with ophthalmic manifestations, type I [juvenile type], not stated as uncontrolled, Diabetes with ophthalmic manifestations, type II or unspecified type, uncontrolled, Diabetes with ophthalmic manifestations, type I [juvenile type], uncontrolled, Diabetes with neurological manifestations, type II or unspecified type, not stated as uncontrolled, Diabetes with neurological manifestations, type I [juvenile type], not stated as uncontrolled, Diabetes with neurological manifestations, type II or unspecified type, uncontrolled, Diabetes with neurological manifestations, type I [juvenile type], uncontrolled, Diabetes with peripheral circulatory disorders, type II or unspecified type, not stated as uncontrolled, Diabetes with peripheral circulatory disorders, type I [juvenile type], not stated as uncontrolled, Diabetes with peripheral circulatory disorders, type II or unspecified type, uncontrolled, Diabetes with peripheral circulatory disorders, type I [juvenile type], uncontrolled, Diabetes with other specified manifestations, type II or unspecified type, not stated as uncontrolled, Diabetes with other specified manifestations, type I [juvenile type], not stated as uncontrolled, Diabetes with other specified manifestations, type II or unspecified type, uncontrolled, Diabetes with other specified manifestations, type I [juvenile type], uncontrolled, Diabetes with unspecified complication, type II or unspecified type, not stated as uncontrolled, Diabetes with unspecified complication, type I [juvenile type], not stated as uncontrolled, Diabetes with unspecified complication, type II or unspecified type, uncontrolled, Diabetes with unspecified complication, type I [juvenile type], uncontrolled, Polyneuropathy in diabetes, Background diabetic retinopathy, Proliferative diabetic retinopathy, Nonproliferative diabetic retinopathy NOS, Mild nonproliferative diabetic retinopathy, Moderate nonproliferative diabetic retinopathy, Severe nonproliferative diabetic retinopathy, Diabetic macular edema, Diabetic cataract, Acetonuria, Diabetes Mellitus Due To Underlying Condition With Hyperosmolarity without nonketotic hyperglycemichyperosmolar coma (NKHHC), Diabetes Mellitus Due To Underlying Condition With Hyperosmolarity With Coma, Diabetes Mellitus Due To Underlying Condition With Ketoacidosis Without Coma, Diabetes Mellitus Due To Underlying Condition With Ketoacidosis With Coma, Diabetes Mellitus Due To Underlying Condition With Diabetic Nephropathy, Diabetes Mellitus Due To Underlying Condition With Diabetic chronic kidney disease, Diabetes |

| | | | |
|---|---|---|---|
| | | E08.65,<br>E08.69, E08.8,<br>E08.9, E08.90,<br>E09.00,<br>E09.01,<br>E09.10,<br>E09.11,<br>E09.21,<br>E09.22,<br>E09.29,<br>E09.311,<br>E09.319,<br>E09.321,<br>E09.329,<br>E09.331,<br>E09.339,<br>E09.341,<br>E09.349,<br>E09.351,<br>E09.359,<br>E09.36,<br>E09.39,<br>E09.40,<br>E09.41,<br>E09.42,<br>E09.43,<br>E09.44,<br>E09.49,<br>E09.51,<br>E09.52,<br>E09.59,<br>E09.610,<br>E09.618,<br>E09.620,<br>E09.621,<br>E09.622,<br>E09.628,<br>E09.630,<br>E09.638, | Mellitus Due to underlying condition with other diabetic kidney complication, Diabetes Mellitus Due To Underlying Condition With Unspecified Diabetic Retinopathy With Macular Edema, Diabetes Mellitus Due To Underlying Condition With Unspecified Diabetic Retinopathy Without Macular Edema, Diabetes Mellitus due to underlying condition with mild nonproliferative diabetic retinopathy with macular edema, Diabetes mellitus due to underlying condition with mild nonproliferative diabetic retinopathy without macular edema, Diabetes mellitus due to underlying condition with moderate nonproliferative diabetic retinopathy with macular edema, Diabetes mellitus due to underlying condition with moderate nonproliferative diabetic retinopathy without macular edema, Diabetes mellitus due to underlying condition with severe nonproliferative diabetic retinopathy with macular edema, Diabetes mellitus due to underlying condition with severe nonproliferative diabetic retinopathy without macular edema, Diabetes mellitus due to underlying condition with proliferative diabetic retinopathy with macular edema, Diabetes mellitus due to underlying condition with proliferative diabetic retinopathy without macular edema, Diabetes Mellitus Due To Underlying Condition With Diabetic Cataract, Diabetes Mellitus Due To Underlying Condition With Other Diabetic Ophthalmic Complication, Diabetes Mellitus Due To Underlying Condition With Diabetic Neuropathy, Unspecified, Diabetes Mellitus Due To Underlying Condition With Diabetic Mononeuropathy, Diabetes Mellitus Due To Underlying Condition With Diabetic Polyneuropathy, Diabetes Mellitus Due To Underlying Condition With Diabetic Autonomic (Poly)Neuropathy, Diabetes Mellitus Due To Underlying Condition With Diabetic Amyotrophy, Diabetes Mellitus Due To Underlying Condition With Other Diabetic Neurological Complication, Diabetes Mellitus Due To Underlying Condition With Diabetic Peripheral Angiopathy Without Gangrene, Diabetes Mellitus Due To Underlying Condition With Diabetic Neuropathic Arthropathy, Diabetes Mellitus Due To Underlying Condition with diabetic neuropathic arthropathy, Diabetes Mellitus Due To Underlying Condition With Diabetic arthropathy, Diabetes Mellitus Due To Underlying Condition With diabetic dermatitis, Diabetes Mellitus Due To Underlying Condition With Foot Ulcer, Diabetes Mellitus Due To Underlying Condition With Other Skin ulcer, Diabetes Mellitus Due To Underlying Condition With other skin complications, Diabetes Mellitus Due To Underlying Condition With periodontal disease, Diabetes Mellitus Due To Underlying Condition With other oral complications, Diabetes Mellitus Due To Underlying Condition With hypoglycemia with coma, Diabetes mellitus due to underlying condition with hypoglycemia without coma, Diabetes Mellitus Due To Underlying Condition With Hyperglycemia, Diabetes Mellitus Due To Underlying Condition With Other Specified Complication, Diabetes Mellitus Due To Underlying Condition With Unspecified Complications, Diabetes mellitus due to underlying condition without Complications, Diabetes Mellitus Due To Underlying Condition Without |

| | | | |
|---|---|---|---|
| | | E09.641, E09.649, E09.65, E09.69, E09.8, E09.9, E10.10, E10.11, E10.21, E10.22, E10.29, E10.311, E10.319, E10.321, E10.329, E10.331, E10.339, E10.341, E10.349, E10.351, E10.359, E10.36, E10.39, E10.40, E10.41, E10.42, E10.43, E10.44, E10.49, E10.51, E10.52, E10.59, E10.610, E10.618, E10.620, E10.621, E10.622, E10.628, E10.630, E10.638, E10.641, | Complications, Drug or Chemical induced diabetes mellitus with hyperosmolarity without nonketotic hyperglycemic-hyperosmolar coma (NKHHC), Drug Or Chemical Induced Diabetes Mellitus With Hyperosmolarity With Coma, Drug Or Chemical Induced Diabetes Mellitus With Ketoacidosis Without Coma, Drug Or Chemical Induced Diabetes, Mellitus With Ketoacidosis With Coma, Drug Or Chemical Induced Diabetes Mellitus With Diabetic Nephropathy, Drug Or Chemical Induced Diabetes Mellitus With diabetic chronic kidney disease, Drug or chemical induced diabetes mellitus with other diabetic kidney complication, Drug Or Chemical Induced Diabetes Mellitus With Unspecified Diabetic Retinopathy With Macular Edema, Drug Or Chemical Induced Diabetes Mellitus With Unspecified Diabetic Retinopathy Without Macular Edema, Drug or chemical induced diabetes mellitus with mild nonproliferative diabetic retinopathy with macular edema, Drug or chemical induced diabetes mellitus with mild nonproliferative diabetic retinopathy without macular edema, Drug or chemical induced diabetes mellitus with moderate nonproliferative diabetic retinopathy with macular edema, Drug or chemical induced diabetes mellitus with moderate nonproliferative diabetic retinopathy without macular edema, Drug or chemical induced diabetes mellitus with severe nonproliferative diabetic retinopathy with macular edema, Drug or chemical induced diabetes mellitus with severe nonproliferative diabetic retinopathy without macular edema, Drug or chemical induced diabetes mellitus with proliferative diabetic retinopathy with macular edema, Drug or chemical induced diabetes mellitus with proliferative diabetic retinopathy without macular edema, Drug Or Chemical Induced Diabetes Mellitus With Diabetic Cataract, Drug Or Chemical Induced Diabetes Mellitus With Other Diabetic Ophthalmic Complication, Drug Or Chemical Induced Diabetes Mellitus With Neurological Complications With Diabetic Neuropathy, Unspecified, Drug Or Chemical Induced Diabetes Mellitus With Neurological Complications With Diabetic Mononeuropathy, Drug Or Chemical Induced Diabetes Mellitus With Neurological Complications With Diabetic Polyneuropathy, Drug Or Chemical Induced Diabetes Mellitus With Neurological Complications With Diabetic Autonomic (Poly)Neuropathy, Drug Or Chemical Induced Diabetes Mellitus With Neurological Complications With Diabetic Amyotrophy, Drug Or Chemical Induced Diabetes Mellitus With Neurological Complications With Other Diabetic Neurological Complication, Drug Or Chemical Induced Diabetes Mellitus With Diabetic Peripheral Angiopathy Without Gangrene, Drug Or Chemical Induced Diabetes Mellitus With Diabetic peripheral angiopathy with gangrene, Drug or chemical induced diabetes mellitus with other circulatory complications, Drug Or Chemical Induced Diabetes Mellitus With Diabetic neuropathic Arthropathy, Drug or chemical induced diabetes mellitus with other diabetic arthropathy, Drug Or Chemical Induced Diabetes Mellitus With diabetic dermatitis, Drug Or Chemical Induced Diabetes Mellitus With foot ulcer, |

| | | | |
|---|---|---|---|
| | | E10.649, E10.65, E10.69, E10.8, E10.9, E11.00, E11.01, E11.21, E11.22, E11.29, E11.311, E11.319, E11.321, E11.329, E11.331, E11.339, E11.341, E11.349, E11.351, E11.359, E11.36, E11.39, E11.40, E11.41, E11.42, E11.51, E11.52, E11.59, E11.610, E11.618, E11.620, E11.621, E11.622, E11.628, E11.630, E11.638, E11.641, E11.649, E11.65, E11.69, E11.8, E11.9, E13.00, | Drug Or Chemical Induced Diabetes Mellitus With Other Skin ulcer, Drug Or Chemical Induced Diabetes Mellitus With other skin complications, Drug Or Chemical Induced Diabetes Mellitus With periodontal disease, Drug Or Chemical Induced Diabetes Mellitus With other oral complications, Drug Or Chemical Induced Diabetes Mellitus With Hypoglycemia with coma, Drug Or Chemical Induced Diabetes Mellitus With hypoglycemia without coma, Drug Or Chemical Induced Diabetes Mellitus With Hyperglycemia, Drug Or Chemical Induced Diabetes Mellitus With Other Specified Complication, Drug Or Chemical Induced Diabetes Mellitus With Unspecified Complications, Drug Or Chemical Induced Diabetes Mellitus without complications, Type 1 Diabetes Mellitus With Ketoacidosis Without Coma, Type 1 Diabetes Mellitus With Ketoacidosis With Coma, Type 1 Diabetes Mellitus With Diabetic Nephropathy, Type 1 Diabetes Mellitus With diabetic chronic kidney disease, Type 1 Diabetes Mellitus With other diabetic kidney complication, Type 1 Diabetes Mellitus With Unspecified Diabetic Retinopathy With Macular Edema, Type 1 Diabetes Mellitus With Unspecified Diabetic Retinopathy Without Macular Edema, Type 1 Diabetes mellitus, with mild nonproliferative diabetic retinopathy with macular edema, Type 1 Diabetes mellitus with mild nonproliferative diabetic retinopathy without macular edema, Type 1 diabetes mellitus with moderate nonproliferative diabetic retinopathy with macular edema, Type 1 diabetes mellitus with moderate nonproliferative diabetic retinopathy without macular edema, Type 1 diabetes mellitus with severe nonproliferative diabetic retinopathy with macular edema, Type 1 diabetes mellitus with severe nonproliferative diabetic retinopathy without macular edema, Type 1 diabetes mellitus with proliferative diabetic retinopathy with macular edema, Type 1 diabetes mellitus with proliferative diabetic retinopathy without macular edema, Type 1 Diabetes Mellitus With Diabetic Cataract, Type 1 Diabetes Mellitus With Other Diabetic Ophthalmic Complication, Type 1 Diabetes Mellitus With Diabetic Neuropathy, Unspecified, Type 1 Diabetes Mellitus With diabetic mononeuropathy, Type 1 Diabetes Mellitus With Diabetic polyneuropathy, Type 1 Diabetes mellitus with diabetic autonomic (poly)neuropathy, Type 1 diabetes mellitus with diabetic amyotrophy, Type 1 diabetes mellitus with other diabetic neurological complication, Type 1 Diabetes Mellitus With Diabetic Peripheral Angiopathy Without Gangrene, Type 1 diabetes mellitus with diabetic peripheral angiopathy with gangrene, Type 1 diabetes mellitus with other circulatory complications, Type 1 diabetes mellitus with diabetic neuropathic arthropathy, Type 1 Diabetes Mellitus With Diabetic arthropathy, Type 1 Diabetes Mellitus With diabetic dermatitis, Type 1 Diabetes Mellitus With Other foot ulcer, Type 1 Diabetes Mellitus With Other Skin ulcer, Type 1 Diabetes Mellitus With other skin complications, Type 1 Diabetes Mellitus With periodontal disease, Type 1 Diabetes Mellitus With other oral complications, Type 1 Diabetes Mellitus With Hypoglycemia With Coma, Type 1 Diabetes Mellitus With |

| | | | |
|---|---|---|---|
| | | E13.01,<br>E13.10,<br>E13.11,<br>E13.21,<br>E13.22,<br>E13.29,<br>E13.311,<br>E13.319,<br>E13.321,<br>E13.329,<br>E13.331,<br>E13.339,<br>E13.341,<br>E13.349,<br>E13.351,<br>E13.359,<br>E13.36,<br>E13.39,<br>E13.40,<br>E13.41,<br>E13.42,<br>E13.43,<br>E13.44,<br>E13.49,<br>E13.51,<br>E13.52,<br>E13.59,<br>E13.610,<br>E13.618,<br>E13.620,<br>E13.621,<br>E13.622,<br>E13.628,<br>E13.630,<br>E13.638,<br>E13.641,<br>E13.649,<br>E13.65, | Hypoglycemia without coma, Type 1 Diabetes Mellitus With Hyperglycemia, Type 1 Diabetes Mellitus With Other Specified Complication, Type 1 Diabetes Mellitus With Unspecified Complications, Type 1 Diabetes Mellitus Without Complications, Type 2 Diabetes Mellitus With Hyperosmolarity Without Nonketotic Hyperglycemic-Hyperosmolar Coma (Nkhhc), Type 2 Diabetes Mellitus With Hyperosmolarity With Coma, Type 2 diabetes mellitus with diabetic nephropathy, Type 2 diabetes mellitus with diabetic chronic kidney disease, Type 2 diabetes mellitus with other diabetic kidney complication, Type 2 Diabetes Mellitus With Unspecified Diabetic Retinopathy With Macular Edema, Type 2 Diabetes Mellitus With Unspecified Diabetic Retinopathy Without Macular Edema, Type 2 Diabetes Mellitus With Mild Nonproliferative Diabetic Retinopathy With Macular Edema, Type 2 Diabetes Mellitus With Mild Nonproliferative Diabetic Retinopathy Without Macular Edema, Type 2 Diabetes Mellitus With Moderate Nonproliferative Diabetic Retinopathy With Macular Edema, Type 2 Diabetes Mellitus with moderate nonproliferative diabetic retinopathy Without Macular Edema, Type 2 Diabetes Mellitus With Severe Nonproliferative Diabetic Retinopathy With Macular Edema, Type 2 Diabetes Mellitus With severe nonproliferative Diabetic Retinopathy Without Macular Edema, Type 2 Diabetes Mellitus with proliferative diabetic retinopathy with macular edema, Type 2 Diabetes Mellitus With proliferative diabetic retinopathy without macular edema, Type 2 Diabetes Mellitus With Diabetic Cataract, Type 2 Diabetes Mellitus With Other Diabetic Ophthalmic Complication, Type 2 Diabetes Mellitus With Diabetic Neuropathy, Unspecified, Type 2 Diabetes Mellitus With Diabetic mononeuropathy, Type 2 Diabetes Mellitus with diabetic polyneuropathy, Type 2 Diabetes Mellitus With Diabetic Peripheral Angiopathy Without Gangrene, Type 2 Diabetes Mellitus With diabetic peripheral Angiopathy With Gangrene, Type 2 Diabetes Mellitus with other circulatory complications, Type 2 diabetes, mellitus with diabetic neuropathic arthropathy, Type 2 diabetes mellitus with other diabetic arthropathy, Type 2 Diabetes Mellitus with diabetic dermatitis, Type 2 Diabetes Mellitus With foot ulcer, Type 2 Diabetes Mellitus With Other Skin ulcer, Type 2 Diabetes Mellitus With other skin complications, Type 2 Diabetes Mellitus With periodontal disease, Type 2 Diabetes Mellitus With other oral complications, Type 2 Diabetes Mellitus With Hypoglycemia With Coma, Type 2 Diabetes Mellitus With Hypoglycemia without coma, Type 2 Diabetes Mellitus With Hyperglycemia, Type 2 Diabetes Mellitus With Other Specified Complication, Type 2 Diabetes Mellitus With Unspecified Complications, Type 2 Diabetes Mellitus Without Complications, Other specified diabetes mellitus with hyperosmolarity without nonketotic hyperglycemic-hyperosmolar coma (NKHHC), Other specified diabetes mellitus with hyperosmolarity with coma, Other specified diabetes mellitus with ketoacidosis without coma, Other specified diabetes mellitus with ketoacidosis with coma, Other specified diabetes mellitus with diabetic |

| | | | |
|---|---|---|---|
| | | E13.69, E13.8, E13.9, R82.4 | nephropathy, Other specified diabetes mellitus with diabetic chronic kidney disease, Other specified diabetes mellitus with other diabetic kidney complication, Other specified diabetes mellitus with unspecified diabetic retinopathy with macular edema, Other specified diabetes mellitus with unspecified diabetic retinopathy without macular edema, Other specified diabetes mellitus with mild nonproliferative diabetic retinopathy with macular edema, Other specified diabetes mellitus with mild nonproliferative diabetic retinopathy without macular edema, Other specified diabetes mellitus with moderate nonproliferative diabetic retinopathy with macular edema, Other specified diabetes mellitus with moderate nonproliferative diabetic retinopathy without macular edema, Other specified diabetes mellitus with severe nonproliferative diabetic retinopathy with macular edema, Other specified diabetes mellitus with severe nonproliferative diabetic retinopathy without macular edema, Other specified diabetes mellitus with proliferative diabetic retinopathy with macular edema, Other specified diabetes mellitus with proliferative diabetic retinopathy without macular edema, Other Specified Diabetes Mellitus With diabetic cataract, Other Specified Diabetes Mellitus With other diabetic ophthalmic complication, Other Specified Diabetes Mellitus With Diabetic neuropathy, unspecified, Other Specified Diabetes Mellitus With Diabetic mononeuropathy, Other Specified Diabetes Mellitus With Diabetic Polyneuropathy, Other Specified Diabetes Mellitus With Diabetic Autonomic (Poly)Neuropathy, Other Specified Diabetes Mellitus With Diabetic Amyotrophy, Other Specified Diabetes Mellitus With Other Diabetic Neurological Complication, Other specified diabetes mellitus with diabetic peripheral angiopathy without gangrene, Other specified diabetes mellitus with diabetic peripheral angiopathy with gangrene, Other Specified Diabetes Mellitus With Other Circulatory Complications, Other specified diabetes mellitus with diabetic neuropathic arthropathy, Other specified diabetes mellitus with other diabetic arthropathy, Other Specified Diabetes Mellitus With Diabetic Dermatitis, Other Specified Diabetes Mellitus With Foot Ulcer, Other Specified Diabetes Mellitus With Other Skin Ulcer, Other specified diabetes mellitus with other skin complications, Other specified diabetes mellitus with periodontal disease, Other specified diabetes mellitus with other oral complications, Other Specified Diabetes Mellitus With Hypoglycemia With Coma, Other Specified Diabetes Mellitus With Hypoglycemia Without Coma, Other Specified Diabetes Mellitus With Hyperglycemia, Other Specified Diabetes Mellitus With Other Specified Complication, Other Specified Diabetes Mellitus With Unspecified Complications, Other Specified Diabetes Mellitus Without Complications, Acetonuria |
| Hypertension | ICD-9, ICD-10 | 401, 401.1, 401.9, 402, 402.01, 402.1, | Malignant essential hypertension, Benign essential hypertension, Unspecified essential hypertension, Malignant hypertensive heart disease without heart failure, Malignant hypertensive heart disease with heart failure, Benign hypertensive heart disease without |

| | | | |
|---|---|---|---|
| | | 402.11, 402.9, 402.91, 403, 403.01, 403.1, 403.11, 403.9, 403.91, 404, 404.01, 404.02, 404.03, 404.1, 404.11, 404.12, 404.13, 404.9, 404.91, 404.92, 404.93, 405.01, 405.09, 405.11, 405.19, 405.91, 405.99, 437.2, 796.2, I10, I11.0, I11.9, I12.0, I12.9, I13.0, I13.10, I13.11, I13.2, I15.0, I15.1, I15.2, I15.8, I15.9 | heart failure, Benign hypertensive heart disease with heart failure, Unspecified hypertensive heart disease without heart failure, Unspecified hypertensive heart disease with heart failure, Hypertensive chronic kidney disease, malignant, with chronic kidney disease stage I through stage IV, or unspecified, Hypertensive chronic kidney disease, malignant, with chronic kidney disease stage V or end stage renal disease, Hypertensive chronic kidney disease, benign, with chronic kidney disease stage I through stage IV, or unspecified, Hypertensive chronic kidney disease, benign, with chronic kidney disease stage V or end stage renal disease, Hypertensive chronic kidney disease, unspecified, with chronic kidney disease stage I through stage IV, or unspecified, Hypertensive chronic kidney disease, unspecified, with chronic kidney disease stage V or end stage renal disease, Hypertensive heart and chronic kidney disease, malignant, without heart failure and with chronic kidney disease stage I through stage IV, or unspecified, Hypertensive heart and chronic kidney disease, malignant, with heart failure and with chronic kidney disease stage I through stage IV, or unspecified, Hypertensive heart and chronic kidney disease, malignant, without heart failure and with chronic kidney disease stage V or end stage renal disease, Hypertensive heart and chronic kidney disease, malignant, with heart failure and with chronic kidney disease stage V or end stage renal disease, Hypertensive heart and chronic kidney disease, benign, without heart failure and with chronic kidney disease stage I through stage IV, or unspecified, Hypertensive heart and chronic kidney disease, benign, with heart failure and with chronic kidney disease stage I through stage IV, or unspecified, Hypertensive heart and chronic kidney disease, benign, without heart failure and with chronic kidney disease stage V or end stage renal disease, Hypertensive heart and chronic kidney disease, benign, with heart failure and chronic kidney disease stage V or end stage renal disease, Hypertensive heart and chronic kidney disease, unspecified, without heart failure and with chronic kidney disease stage I through stage IV, or unspecified, Hypertensive heart and chronic kidney disease, unspecified, with heart failure and with chronic kidney disease stage I through stage IV, or unspecified, Hypertensive heart and chronic kidney disease, unspecified, without heart failure and with chronic kidney disease stage V or end stage renal disease, Hypertensive heart and chronic kidney disease, unspecified, with heart failure and chronic kidney disease stage V or end stage renal disease, Malignant renovascular hypertension, Other malignant secondary hypertension, Benign renovascular hypertension, Other benign secondary hypertension, Unspecified renovascular hypertension, Other unspecified secondary hypertension, Hypertensive encephalopathy, Elevated blood pressure reading without diagnosis of hypertension, Essential (Primary) Hypertension, Hypertensive Heart Disease with Heart Failure, Hypertensive Heart Disease without Heart Failure, Hypertensive Chronic Kidney Disease with Stage 5 Chronic Kidney Disease or end stage renal disease, Hypertensive Chronic Kidney |

| | | | |
|---|---|---|---|
| | | | Disease with Stage 1 through Stage 4 Chronic Kidney Disease, or unspecified chronic kidney disease, Hypertensive heart and chronic kidney disease with heart failure and stage 1 through stage 4 chronic kidney disease, or unspecified chronic kidney disease , Hypertensive heart and chronic kidney disease without heart failure, with stage 1 through stage 4 chronic kidney disease, or unspecified chronic kidney disease , Hypertensive heart and chronic kidney disease without heart failure, with stage 5 chronic kidney disease, or end stage renal disease, Hypertensive heart and chronic kidney disease with heart failure and with stage 5 chronic kidney disease, or end stage renal disease, Renovascular Hypertension, Hypertension secondary to other renal disorders, Hypertension secondary to endocrine disorders, Other Secondary Hypertension, Secondary hypertension, unspecified, Endovascular replacement of aortic valve, Transapical replacement of aortic valve, Open heart valvuloplasty without replacement, unspecified valve, Open heart valvuloplasty of aortic valve without replacement, Open heart valvuloplasty of mitral valve without replacement, Open heart valvuloplasty of pulmonary valve without replacement, Open heart valvuloplasty of tricuspid valve without replacement, Open and other replacement of unspecified heart valve, Open and other replacement of aortic valve with tissue graft, Open and other replacement of aortic valve, Open and other replacement of mitral valve with tissue graft, Open and other replacement of mitral valve, Open and other replacement of pulmonary valve with tissue graft, Open and other replacement of pulmonary valve, Open and other replacement of tricuspid valve with tissue graft, Open and other replacement of tricuspid valve, Percutaneous balloon valvuloplasty, Mitral stenosis, Rheumatic Mitral Insufficiency, Mitral stenosis with insufficiency, Other unspecified mitral valve disease, Mitral valve stenosis and aortic valve stenosis, Mitral valve stenosis and aortic valve insufficiency, Mitral valve insufficiency and aortic valve stenosis, Mitral valve insufficiency and aortic valve insufficiency, Multiple involvement of mitral and aortic valves, Mitral and aortic valve diseases, unspecified, Heart valve replaced by transplant, Heart valve replaced by other means |
| Hyperlipidemia | ICD-9, ICD-10 | 272, 272.4, 272.2, 272.3, 272.6, 759.9, 272.7, 272.1, 272.5, 272.8, 272.9, E78.0, E78.1, E78.2, E78.3, E78.4, E78.5, E78.7, E88.1, E78.81, | Pure hypercholesterolemia, Other hyperlipidemia, Mixed hyperlipidemia, Hyperchylomicronemia, Lipoprotein deficiency, Disorders of bile acid and cholesterol metabolism, Lipidoses, Pure hyperglyceridemia, Hyperlipidemia, unspecified, Other specified metabolic disorders, Disorder of lipoprotein metabolism, unspecified, Pure hypercholesterolemia, Pure hyperglyceridemia, Mixed hyperlipidemia, Hyperchylomicronemia, Other hyperlipidemia, Hyperlipidemia, unspecified, Disorders of bile acid and cholesterol metabolism, Lipodystrophy, Lipid dermatoarthritis, Lipidoses, Other specified metabolic disorders, Lipidoses, Lipidoses, Lipidoses, Other lipoprotein metabolism disorders, Lipoprotein deficiency, Disorder of lipoprotein metabolism, unspecified, Disorder of fatty-acid metabolism, unspecified, Other lipid |

| | | | |
|---|---|---|---|
| | | E77.1, E88.89, E75.21, E75.22, E77.0, E78.89, E78.6, E78.9, E71.30, E75.5, E78.79, E75.6, E78.70 | storage disorders, Other disorders of bile acid and cholesterol metabolism, Lipid storage disorder, unspecified, Disorder of bile acid and cholesterol metabolism, unspecified |
| Obesity | ICD-9; ICD-10 | 278.00, 278.01; E66.1, E66.01, E66.09, E66.9, E66.2, I66.8 | Obesity unspecified, Morbid obesity;<br>Drug-induced obesity, Morbid obesity due to access calories,<br>Other obesity due to access calories, Obesity unspecified, Morbid obesity with alveolar hypoventilation, Other obesity |
| Myocardial infarction | ICD-9; ICD-10 | 410.xx; I21.0x-I21.3x, I21.4, I22.0x-I22.1x, I22.2x, I22.8x-I22.9x, I23.xx, I24.1, I25.2 | Acute myocardial infarction of xx wall;<br>ST elevation (STEMI) myocardial infarction involving xx artery, Non-ST elevation (NSTEMI) myocardial infarction, Subsequent ST elevation (STEMI) myocardial infarction of anterior wall, Subsequent non-ST elevation (NSTEMI) myocardial infarction, Subsequent ST elevation (STEMI) myocardial infarction of other sites, xx complications following acute myocardial infarction, Dressler's syndrome, Old myocardial infarction |
| Heart failure | ICD-9, ICD-10 | 402.xx, 404.xx, 428.1, 428.2x, 428.3x, 428.4x, 428.9, I11.0, I13.x, I50.1, I50.2x, I50.3x, I50.4x, I50.9 | xx hypertensive heart disease with heart failure, Hypertensive heart and chronic kidney disease, malignant, with heart failure and with chronic kidney disease stage xx, Left heart failure, Systolic heart failure, Diastolic heart failure, Combined systolic and diastolic heart failure, Heart failure- unspecified, Hypertensive heart disease with heart failure, Hypertensive heart and chronic kidney disease with heart failure and stage xx, Left ventricular failure, unspecified, Systolic (congestive) heart failure, Diastolic (congestive) heart failure, Combined systolic (congestive) and diastolic (congestive) heart failure, Heart failure - unspecified |
| Vascular dissection | ICD-9, ICD-10 | 443.21, 443.22, 443.23, 443.24, 443.29, 441.00, 441.01, 441.02, 441.03, I25.42, I77.71, I77.72, I77.73, I77.74, I77.79, I71.00, | Dissection of carotid artery, Dissection of iliac artery, Dissection of renal artery, Dissection of vertebral artery, Dissection of other artery, Dissection of unspecified site of aorta, Dissection of thoracic aorta, Dissection of abdominal aorta, Dissection of thoracoabdominal aorta, Coronary artery dissection, Dissection of carotid artery, Dissection of iliac artery, Dissection of renal artery, Dissection of vertebral artery, Dissection of other artery, Dissection of unspecified site of aorta, Dissection of thoracic aorta, Dissection of abdominal aorta, Dissection of thoracoabdominal aorta |

| | | | |
|---|---|---|---|
| | | I71.01, I71.02, I71.03 | |
| Venous thromboembolism | ICD-9, ICD-10 | I82.Axx, I82.Bxx, I82.Cxx, Z86.718, 453, 453.1, 453.2, 453.3, 453.4x, 453.5x, 453.6, 453.7x, 453.8x, 453.9, 557, A82.A22, I82.21x, I82.22x, I82.29x, I82.3, I82.40x, I82.41x, I82.42x, I82.43x, I82.44x, I82.49x, I82.4Yx, I82.4Zx, I82.50x, I82.51x, I82.52x, I82.53x, I82.54x, i82.59x, I82.5Yx, I82.5Zx, I82.6xx, I82.7xx, I82.89x, | Chronic embolism and thrombosis of axillary vein, Acute/Chronic embolism and thrombosis of xx subclavian vein, Acute embolism and thrombosis of xx jugular vein, Personal history of other venous thrombosis and embolism, Budd-chiari syndrome, Thrombophlebitis migrans, Other venous embolism and thrombosis of inferior vena cava, Other venous embolism and thrombosis of renal vein, Acute venous embolism and thrombosis of deep vessels of lower extremity, Chronic venous embolism and thrombosis of deep vessels of lower extremity, Venous embolism and thrombosis of superficial vessels of lower extremity, Chronic venous embolism and thrombosis of deep veins of upper extremity, Acute venous embolism and thrombosis of deep veins of upper extremity, Other venous embolism and thrombosis of unspecified site, Acute vascular insufficiency of intestine, Chronic embolism and thrombosis of left axillary vein, Embolism and thrombosis of superior vena cava, Embolism and thrombosis of inferior vena cava, Embolism and thrombosis of other thoracic veins, Embolism and thrombosis of renal vein, Acute embolism and thrombosis of unspecified deep veins of lower extremity, Acute embolism and thrombosis of femoral vein, Acute embolism and thrombosis of iliac vein, Acute embolism and thrombosis of popliteal vein, Acute embolism and thrombosis of tibial vein, Acute embolism and thrombosis of other specified deep vein of lower extremity, Acute embolism and thrombosis of unspecified deep veins of proximal lower extremity, Acute embolism and thrombosis of unspecified deep veins of distal lower extremity, Chronic embolism and thrombosis of unspecified deep veins of lower extremity, Chronic embolism and thrombosis of femoral vein, Chronic embolism and thrombosis of iliac vein, Chronic embolism and thrombosis of popliteal vein, Chronic embolism and thrombosis of tibial vein, Chronic embolism and thrombosis of other specified deep vein of lower extremity, Chronic embolism and thrombosis of unspecified deep veins of proximal lower extremity, Chronic embolism and thrombosis of unspecified deep veins of distal lower extremity, Acute embolism and thrombosis of deep veins of upper extremity, Chronic embolism and thrombosis of unspecified veins of upper extremity, Embolism and thrombosis of other specified veins, Embolism and thrombosis of unspecified vein, Acute embolism and thrombosis of axillary vein |

| | | | |
|---|---|---|---|
| | | I82.9x, I82.Axx | |
| Pulmonary embolism | ICD-9, ICD-10 | 415.11, 415.12, 415.13, 415.19, 673.8x, I26.01, I26.02, I26.09, I26.90, I26.92, I26.99, I27.88, V12.55, Z86.711 | Iatrogenic pulmonary embolism and infarction, Septic pulmonary embolism, Saddle embolus of pulmonary artery, Other pulmonary embolism and infarction, Septic pulmonary embolism with acute cor pulmonate, Saddle embolus of pulmonary artery with acute cor pulmonate, Other pulmonary embolism with acute cor pulmonale, Septic pulmonary embolism without acute cor pulmonale, Saddle embolus of pulmonary artery without acute cor pulmonale, Other pulmonary embolism without acute cor pulmonale, Chronic pulmonary embolism, Personal history of pulmonary embolism, Personal history of pulmonary embolism |
| Systemic embolism | ICD-9, ICD-10 | 444.xx, I74.xx, I76 | Embolism and thrombosis of xx, Embolism and thrombosis of xx, Septic arterial embolism |
| Transient ischemic attack | ICD-9, ICD-10 | 362.31, 362.32, 362.33, 362.34, 388.02, 430, 431, 432.9, 433.01, 433.11, 433.21, 433.31, 433.81, 433.91, 434, 434.01, 434.1, 434.11, 434.9, 434.91, 435, 435.1, 435.2, 435.3, 435.8, 435.9, 437.1, 437.7, 437.9, 438.1, 438.11, 438.12, 438.13, 438.14, 438.2, 438.21, 438.22, 438.81, 438.82, 438.83, 438.89, 438.9, 997.02, | Central retinal artery occlusion, Retinal arterial branch occlusion, Partial retinal arterial occlusion, Transient retinal arterial occlusion, Transient ischemic deafness, Subarachnoid hemorrhage, Intracerebral hemorrhage, Unspecified intracranial hemorrhage, Occlusion and stenosis of basilar artery with cerebral infarction, Occlusion and stenosis of carotid artery with cerebral infarction, Occlusion and stenosis of vertebral artery with cerebral infarction, Occlusion and stenosis of multiple and bilateral precerebral arteries with cerebral infarction, Occlusion and stenosis of other specified precerebral artery with cerebral infarction, Occlusion and stenosis of unspecified precerebral artery with cerebral infarction, Cerebral thrombosis without mention of cerebral infarction, Cerebral thrombosis with cerebral infarction, Cerebral embolism without mention of cerebral infarction, Cerebral embolism with cerebral infarction, Cerebral artery occlusion, unspecified without mention of cerebral infarction, Cerebral artery occlusion, unspecified with cerebral infarction, Basilar artery syndrome, Vertebral artery syndrome , Subclavian steal syndrome, Vertebrobasilar artery syndrome, Other specified transient cerebral ischemias, Unspecified transient cerebral ischemia, Other generalized ischemic cerebrovascular disease, Transient global amnesia, Unspecified cerebrovascular disease, Late effects of cerebrovascular disease, speech and language deficit, unspecified, Late effects of cerebrovascular disease, aphasia, Late effects of cerebrovascular disease, dysphasia, Late effects of cerebrovascular disease, dysarthria, Late effects of cerebrovascular disease, fluency disorder, Late effects of cerebrovascular disease, hemiplegia affecting unspecified side, Late effects of cerebrovascular disease, hemiplegia affecting |

| | | | |
|---|---|---|---|
| | | V12.54, G45.0, G45.1, G45.2, G45.3, G45.4, G45.8, G46.3, G46.4, H34.00, H34.01, H34.02, H34.03, H34.10, H34.11, H34.12, H34.13, H34.211, H34.212, H34.213, H34.219, H34.231, H34.232, H34.233, H34.239, H93.099, I60.9, I61.9, I62.9, I63.00, I63.011, I63.012, I63.019, I63.111, I63.112, I63.119, I63.12, I63.131, I63.132, I63.139, I63.19, I63.20, I63.211, I63.212, I63.219, I63.22, I63.231, | dominant side, Late effects of cerebrovascular disease, hemiplegia affecting nondominant side, Other late effects of cerebrovascular disease, apraxia, Other late effects of cerebrovascular disease, dysphagia, Other late effects of cerebrovascular disease, facial weakness, Other late effects of cerebrovascular disease, Unspecified late effects of cerebrovascular disease, Iatrogenic cerebrovascular infarction or hemorrhage, Personal history of transient ischemic attack (TIA), and cerebral infarction without residual deficits, Vertebro-Basilar Artery Syndrome, Carotid Artery Syndrome, Multiple and bilateral precerebral artery syndromes, Amaurosis fugax, Transient Global Amnesia, Other transient cerebral ischemic attacks and related syndromes, Brain stem stroke syndrome, Cerebellar stroke syndrome, Transient Retinal Artery Occlusion, Unspecified Eye, Transient retinal artery occlusion, right eye, Transient retinal artery occlusion, left eye, Transient retinal artery occlusion, bilateral, Central retinal artery occlusion, unspecified eye, Central retinal artery occlusion, right eye, Central retinal artery occlusion, left eye, Central retinal artery occlusion, bilateral, Partial retinal artery occlusion, right eye, Partial retinal artery occlusion, left eye, Partial retinal artery occlusion, bilateral, Partial Retinal Artery Occlusion, Unspecified Eye, Retinal artery branch occlusion, right eye, Retinal artery branch occlusion, left eye, Retinal artery branch occlusion, bilateral, Retinal Artery Branch Occlusion, Unspecified Eye, Unspecified Degenerative and Vascular Disorders of Unspecified Ear, Nontraumatic Subarachnoid Hemorrhage, Unspecified, Nontraumatic intracerebral Hemorrhage, unspecified, Nontraumatic Intracranial Hemorrhage, Unspecified, Cerebral infarction due to thrombosis of unspecified precerebral artery, Cerebral infarction due to thrombosis of right vertebral artery, Cerebral infarction due to thrombosis of left vertebral artery, Cerebral infarction due to thrombosis of unspecified vertebral artery, Cerebral infarction due to embolism of right vertebral artery, Cerebral infarction due to embolism of left vertebral artery, Cerebral infarction due to embolism of unspecified vertebral artery , Cerebral infarction due to embolism of basilar artery, Cerebral infarction due to embolism of right carotid artery, Cerebral infarction due to embolism of left carotid artery, Cerebral infarction due to embolism of unspecified carotid artery, Cerebral infarction due to embolism of other precerebral artery, Cerebral infarction due to unspecified occlusion or stenosis of unspecified precerebral arteries , Cerebral infarction due to unspecified occlusion or stenosis of right vertebral arteries, Cerebral infarction due to unspecified occlusion or stenosis of left vertebral arteries, Cerebral infarction due to unspecified occlusion or stenosis of unspecified vertebral arteries, Cerebral Infraction Due to Unspecified Occlusion or Stenosis of Basilar Arteries, Cerebral infarction due to unspecified occlusion or stenosis of right carotid arteries, Cerebral infarction due to unspecified occlusion or stenosis of left carotid arteries, Cerebral infarction due to unspecified occlusion or stenosis of unspecified carotid arteries, Cerebral infarction due to unspecified occlusion or stenosis of other |

| | | | |
|---|---|---|---|
| | | I63.232, I63.239, I63.29, I63.30, I63.311, I63.312, I63.319, I63.321, I63.322, I63.329, I63.331, I63.332, I63.339, I63.341, I63.342, I63.349, I63.40, I63.411, I63.412, I63.419, I63.421, I63.422, I63.429, I63.431, I63.432, I63.439, I63.49, I63.50, I63.511, I63.512, I63.519, I63.521, I63.522, I63.529, I63.531, I63.532, I63.539, I63.541, I63.542, I63.549, I63.59, I63.6, | precerebral arteries, Cerebral infarction due to thrombosis of unspecified cerebral artery , Cerebral infarction due to thrombosis of right middle cerebral artery, Cerebral infarction due to thrombosis of left middle cerebral artery, Cerebral infarction due to thrombosis of unspecified middle cerebral artery, Cerebral infarction due to thrombosis of right anterior cerebral artery, Cerebral infarction due to thrombosis of left anterior cerebral artery, Cerebral infarction due to thrombosis of unspecified anterior cerebral artery, Cerebral infarction due to thrombosis of right posterior cerebral artery, Cerebral infarction due to thrombosis of left posterior cerebral artery, Cerebral infarction due to thrombosis of unspecified posterior cerebral artery, Cerebral infarction due to thrombosis of right cerebellar artery, Cerebral infarction due to thrombosis of left cerebellar artery, Cerebral infarction due to thrombosis of unspecified cerebellar artery, Cerebral infarction due to embolism of unspecified cerebral artery , Cerebral infarction due to embolism of right middle cerebral artery, Cerebral infarction due to embolism of left middle cerebral artery, Cerebral infarction due to embolism of unspecified middle cerebral artery, Cerebral infarction due to embolism of right anterior cerebral artery, Cerebral infarction due to embolism of left anterior cerebral artery, Cerebral infarction due to embolism of unspecified anterior cerebral artery, Cerebral infarction due to embolism of right posterior cerebral artery, Cerebral infarction due to embolism of left posterior cerebral artery, Cerebral infarction due to embolism of unspecified posterior cerebral artery , Cerebral infarction due to embolism of other cerebral artery, Cerebral infarction due to unspecified occlusion or stenosis of unspecified cerebral artery , Cerebral infarction due to unspecified occlusion or stenosis of right middle cerebral artery, Cerebral infarction due to unspecified occlusion or stenosis of left middle cerebral artery, Cerebral infarction due to unspecified occlusion or stenosis of unspecified middle cerebral artery, Cerebral infarction due to unspecified occlusion or stenosis of right anterior cerebral artery, Cerebral infarction due to unspecified occlusion or stenosis of left anterior cerebral artery, Cerebral infarction due to unspecified occlusion or stenosis of unspecified anterior cerebral artery, Cerebral infarction due to unspecified occlusion or stenosis of right posterior cerebral artery, Cerebral infarction due to unspecified occlusion or stenosis of left posterior cerebral artery, Cerebral infarction due to unspecified occlusion or stenosis of unspecified posterior cerebral artery, Cerebral infarction due to unspecified occlusion or stenosis of right cerebellar artery, Cerebral infarction due to unspecified occlusion or stenosis of left cerebellar artery, Cerebral infarction due to unspecified occlusion or stenosis of unspecified cerebellar artery, Cerebral infarction due to unspecified occlusion or stenosis of other cerebral artery , Cerebral infarction due to cerebral venous thrombosis, nonpyogenic, Other cerebral infarction, Cerebral infarction, unspecified, Occlusion and stenosis of right middle cerebral artery, Occlusion and stenosis of left middle cerebral artery, Occlusion and stenosis of bilateral middle cerebral arteries, |

| | | | |
|---|---|---|---|
| | | I63.8, I63.9, I66.01, I66.02, I66.03, I66.09, I66.11, I66.12, I66.13, I66.19, I66.21, I66.22, I66.23, I66.29, I66.3, I66.8, I66.9, I67.81, I67.82, I67.841, I67.848, I67.89, I67.9, I69.80, I69.81, I69.820, I69.821, I69.822, I69.823, I69.828, I69.831, I69.832, I69.833, I69.834, I69.839, I69.841, I69.842, I69.843, I69.844, I69.849, I69.851, I69.852, I69.853, I69.854, I69.859, I69.861, I69.862, I69.863, I69.864, I69.865, | Occlusion and stenosis of unspecified middle cerebral artery, Occlusion and stenosis of right anterior cerebral artery, Occlusion and stenosis of left anterior cerebral artery, Occlusion and stenosis of bilateral anterior cerebral arteries, Occlusion and stenosis of unspecified anterior cerebral artery, Occlusion and stenosis of right posterior cerebral artery, Occlusion and stenosis of left posterior cerebral artery, Occlusion and stenosis of bilateral posterior cerebral arteries, Occlusion and stenosis of unspecified posterior cerebral artery , Occlusion and stenosis of cerebellar arteries, Occlusion and stenosis of other cerebral arteries, Occlusion and stenosis of unspecified cerebral artery, Acute cerebrovascular insufficiency, Cerebral ischemia, Reversible cerebrovascular vasoconstriction syndrome, Other cerebrovascular vasospasm and vasoconstriction , Other cerebrovascular disease, Cerebrovascular Disease, unspecified, Unspecified sequelae of other cerebrovascular disease, Cognitive deficits following other cerebrovascular disease, Aphasia following other cerebrovascular disease, Dysphasia following other cerebrovascular disease, Dysarthria following other cerebrovascular disease, Fluency disorder following other cerebrovascular disease, Other speech and language deficits following other cerebrovascular disease, Monoplegia of upper limb following other cerebrovascular disease affecting right dominant side, Monoplegia of upper limb following other cerebrovascular disease affecting left dominant side, Monoplegia of upper limb following other cerebrovascular disease affecting right non-dominant side, Monoplegia of upper limb following other cerebrovascular disease affecting left non-dominant side, Monoplegia of upper limb following other cerebrovascular disease affecting unspecified side, Monoplegia of lower limb following other cerebrovascular disease affecting right dominant side, Monoplegia of lower limb following other cerebrovascular disease affecting left dominant side, Monoplegia of lower limb following other cerebrovascular disease affecting right non-dominant side, Monoplegia of lower limb following other cerebrovascular disease affecting left non-dominant side, Monoplegia of lower limb following other cerebrovascular disease affecting unspecified side, Hemiplegia and hemiparesis following other cerebrovascular disease affecting right dominant side, Hemiplegia and hemiparesis following other cerebrovascular disease affecting left dominant side, Hemiplegia and hemiparesis following other cerebrovascular disease affecting right non-dominant side, Hemiplegia and hemiparesis following other cerebrovascular disease affecting left non-dominant side, Hemiplegia and hemiparesis following other cerebrovascular disease affecting unspecified side, Other paralytic syndrome following other cerebrovascular disease affecting right dominant side, Other paralytic syndrome following other cerebrovascular disease affecting left dominant side, Other paralytic syndrome following other cerebrovascular disease affecting right non-dominant side, Other paralytic syndrome following other cerebrovascular disease affecting left non-dominant side, Other paralytic syndrome following other cerebrovascular disease, |

| | | | |
|---|---|---|---|
| | | I69.869, I69.890, I69.891, I69.892, I69.893, I69.898, I69.90, I69.91, I69.920, I69.921, I69.922, I69.923, I69.928, I69.931, I69.932, I69.933, I69.934, I69.939, I69.941, I69.942, I69.943, I69.944, I69.949, I69.951, I69.952, I69.953, I69.954, I69.959, I69.961, I69.962, I69.963, I69.964, I69.965, I69.969, I69.990, I69.991, I69.992, I69.993, I69.998, I97.810, | bilateral, Other paralytic syndrome following other cerebrovascular disease affecting unspecified side, Apraxia following other cerebrovascular disease, Dysphagia following other cerebrovascular disease, Facial weakness following other cerebrovascular disease, Ataxia following other cerebrovascular disease, Other sequelae of other cerebrovascular disease , Unspecified Sequelae of unspecified cerebrovascular disease, Cognitive deficits following unspecified cerebrovascular disease, Aphasia following unspecified cerebrovascular disease, Dysphasia following unspecified cerebrovascular disease , Dysarthria following unspecified Cerebrovascular disease, Fluency disorder following unspecified cerebrovascular disease , Other speech and language deficits following unspecified cerebrovascular disease, Monoplegia of upper limb following unspecified cerebrovascular disease affecting right dominant side, Monoplegia of upper limb following unspecified cerebrovascular disease affecting left dominant side, Monoplegia of upper limb following unspecified cerebrovascular disease affecting right non-dominant side, Monoplegia of upper limb following unspecified cerebrovascular disease affecting left non-dominant side, Monoplegia of upper limb following unspecified cerebrovascular disease affecting unspecified side, Monoplegia of lower limb following unspecified cerebrovascular disease affecting right dominant side, Monoplegia of lower limb following unspecified cerebrovascular disease affecting left dominant side, Monoplegia of lower limb following unspecified cerebrovascular disease affecting right non-dominant side, Monoplegia of lower limb following unspecified cerebrovascular disease affecting left non-dominant side, Monoplegia of lower limb following unspecified cerebrovascular disease affecting unspecified side, Hemiplegia and hemiparesis following unspecified cerebrovascular disease affecting right dominant side , Hemiplegia and hemiparesis following unspecified cerebrovascular disease affecting left dominant side , Hemiplegia and hemiparesis following unspecified cerebrovascular disease affecting right non-dominant side, Hemiplegia and hemiparesis following unspecified cerebrovascular disease affecting left non-dominant side , Hemiplegia and hemiparesis following unspecified cerebrovascular disease affecting unspecified side , Other paralytic syndrome following unspecified cerebrovascular disease affecting right dominant side, Other paralytic syndrome following unspecified cerebrovascular disease affecting left dominant side, Other paralytic syndrome following unspecified cerebrovascular disease affecting right non-dominant side, Other paralytic syndrome following unspecified cerebrovascular disease affecting left non-dominant side, Other paralytic syndrome following unspecified cerebrovascular disease, bilateral, Other paralytic syndrome following unspecified cerebrovascular disease affecting unspecified side, Apraxia following unspecified cerebrovascular disease, Dysphagia following unspecified cerebrovascular disease , Facial weakness following unspecified cerebrovascular disease , Ataxia following unspecified cerebrovascular disease, Other |

| | | | |
|---|---|---|---|
| | | I97.811, I97.820, I97.821, Z86.73 | sequelae following unspecified cerebrovascular disease , Intraoperative Cerebrovascular Infarction During cardiac surgery, Intraoperative cerebrovascular infarction during other surgery , Postprocedural cerebrovascular infarction during cardiac surgery, Postprocedural cerebrovascular infarction during other surgery, Personal history of transient ischemic attack (TIA), and cerebral infarction without residual deficits |
| Hemorrhagic CVA | ICD-9, ICD-10 | 430, 431, I60, I60.0, I60.00, I60.01, I60.02, I60.1, I60.10, I60.11, I60.12, I60.2, I60.20, I60.21, I60.22, I60.3, I60.30, I60.31, I60.32, I60.4, I60.5, I60.50, I60.51, I60.52, I60.6, I60.7, I60.8, I60.9, I61, I61.0, I61.1, I61.2, I61.3, I61.4, I61.5, I61.6, I61.8, I61.9, I62.9 | Subarachnoid hemorrhage, Intracerebral hemorrhage, Nontraumatic subarachnoid hemorrhage, Nontraumatic subarachnoid hemorrhage from carotid siphon and bifurcation, Nontraumatic subarachnoid hemorrhage from unspecified carotid siphon and bifurcation, Nontraumatic subarachnoid hemorrhage from right carotid siphon and bifurcation, Nontraumatic subarachnoid hemorrhage from left carotid siphon and bifurcation, Nontraumatic subarachnoid hemorrhage from middle cerebral artery, Nontraumatic subarachnoid hemorrhage from unspecified middle cerebral artery, Nontraumatic subarachnoid hemorrhage from right middle cerebral artery, Nontraumatic subarachnoid hemorrhage from left middle cerebral artery, Nontraumatic subarachnoid hemorrhage from anterior communicating artery, Nontraumatic subarachnoid hemorrhage from unspecified anterior communicating artery, Nontraumatic subarachnoid hemorrhage from right anterior communicating artery, Nontraumatic subarachnoid hemorrhage from left anterior communicating artery, Nontraumatic subarachnoid hemorrhage from posterior communicating artery, Nontraumatic subarachnoid hemorrhage from unspecified posterior communicating artery, Nontraumatic subarachnoid hemorrhage from right posterior communicating artery, Nontraumatic subarachnoid hemorrhage from left posterior communicating artery, Nontraumatic subarachnoid hemorrhage from basilar artery, Nontraumatic subarachnoid hemorrhage from vertebral artery, Nontraumatic subarachnoid hemorrhage from unspecified vertebral artery, Nontraumatic subarachnoid hemorrhage from right vertebral artery, Nontraumatic subarachnoid hemorrhage from left vertebral artery, Nontraumatic subarachnoid hemorrhage from other intracranial arteries, Nontraumatic subarachnoid hemorrhage from unspecified intracranial artery, Other nontraumatic subarachnoid hemorrhage, Nontraumatic subarachnoid hemorrhage, unspecified, Nontraumatic intracerebral hemorrhage, Nontraumatic intracerebral hemorrhage in hemisphere, subcortical, Nontraumatic intracerebral hemorrhage in hemisphere, cortical, Nontraumatic intracerebral hemorrhage in hemisphere, unspecified, Nontraumatic intracerebral hemorrhage in brain stem, Nontraumatic intracerebral hemorrhage in cerebellum, Nontraumatic intracerebral hemorrhage, intraventricular, Nontraumatic intracerebral hemorrhage, multiple localized, Other nontraumatic intracerebral hemorrhage, Nontraumatic intracerebral hemorrhage, unspecified, Nontraumatic Intracranial Hemorrhage, Unspecified |

| | | | |
|---|---|---|---|
| Ischemic CVA | ICD-9, ICD-10 | I63.xxx, 362.3x, 433.xx, 434.01, 434.11, 434.91, 436 | Cerebral infarction, Central retinal artery occlusion, Occlusion and stenosis of artery with cerebral infarction, Cerebral thrombosis with cerebral infarction, Cerebral embolism with cerebral infarction, Cerebral artery occlusion, unspecified with cerebral infarction, Acute, but ill-defined cerebrovascular disease |
| Supraventricular tachycardia | ICD-9, ICD-10 | 427, 427.2, I47.9, I47.1 | Paroxysmal supraventricular tachycardia, Paroxysmal tachycardia, unspecified, Paroxysmal tachycardia, unspecified, Supraventricular tachycardia |
| Ventricular tachycardia | ICD-9, ICD-10 | 427.69, 427.1, I47.2, I49.40, I49.3 | Ventricular premature beats, Paroxysmal ventricular tachycardia, Ventricular tachycardia, Premature ventricular depolarization, Premature ventricular contraction |
| Cardiac arrest | ICD-9; ICD-10 | 427.5, 427.42, 427.4, 427.41, I49.01, I49.02, I46.9, I49.0 | Sudden cardiac arrest, Ventricular flutter, Ventricular fibrillation or flutter, Ventricular fibrillation, Ventricular fibrillation, Ventricular flutter, Sudden cardiac arrest, Ventricular fibrillation or flutter |

**eTable 3.** Hypertensive disorders of pregnancy definitions

| **Category** | **ICD-9, ICD-10, and CPT codes** |
|---|---|
| Chronic hypertension | 642.0x, 642.8x, 642.9x, O16.x |
| Gestational hypertension | O13.x |
| Preeclampsia | 642.4x, 642.5x, 642.7x, O11.xx, O14.xx |
| Eclampsia | 642.6x, O15.xx |
| HELLP Syndrome | O14.24 |

**eTable 4. Individual-level baseline demographics, clinical, and pregnancy characteristics**

| | **PADME (N=38,997)** |
|---|---|
| Age range, years | 26 - 38 |
| Race/ethnicity | |
| White, n (%) | 23188 (59%) |
| Black, n (%) | 4173 (11%) |
| Asian, n (%) | 2559 (7%) |
| Hispanic ethnicity, n (%) | 4986 (13%) |
| Other, n (%) | 4091 (10%) |
| National ADI score | 23 (12, 43) |
| State ADI score | 4.5 ± 2.1 |
| Body mass index, $kg/m^2$ | 25.8 ± 5.8 |
| Systolic blood pressure, mmHg | 114 ± 12 |
| Diastolic blood pressure, mmHg | 70 ± 9 |
| Pulse, bpm | 78 ± 15 |
| Alcohol use, n (%) | 1766 (4%) |
| Smoking use, n (%) | 1394 (3%) |
| Hemoglobin, g/dL | 12.8 ± 1.0 |
| Pre-existing obesity, n (%) | 3650 (9%) |
| Pre-existing diabetes, n (%) | 978 (2%) |
| Pre-existing hypertension, n (%) | 2648 (7%) |
| Pre-existing hyperlipidemia, n (%) | 3467 (9%) |
| Pre-existing CVD, n (%) | 1333 (3%) |

Individual-level data presented at the start of the first pregnancy captured in PADME. Values are mean ± SD, median (Q1, Q3), or n (%). Values shown exclude missing data. Abbreviations: ADI = area deprivation index, PADME = Predictive Analysis with Deep Learning Models for Maternal Endpoints

**eTable 5. Pregnancy characteristics of PADME**

| | PADME (N = 57,683) | Cardiovascular complication (N = 8,457) | Without cardiovascular complication (N = 49,226) |
|---|---|---|---|
| Live births, n (%) | 52,551 (91%) | 7333 (87%) | 45218 (92%) |
| Mixed births, n (%) | 38 (0.1%) | 8 (0.1%) | 30 (0.1%) |
| Stillbirth, n (%) | 515 (1%) | 142 (2%) | 373 (1%) |
| Unclassified delivery, n (%) | 4579 (8%) | 974 (11%) | 3605 (7%) |
| Gravidity, n | 2.6 ± 1.7 | 2.6 ± 1.8 | 2.6 ± 1.7 |
| Parity, n | 1.3 ± 1.2 | 1.2 ± 1.2 | 1.3 ± 1.2 |
| Estimated gestational age, weeks | 39 ± 4 | 37 ± 4 | 39 ± 3 |

Values are mean ± SD, median (Q1, Q3), or n (%). Values shown exclude missing data. Abbreviations: PADME = Predictive Analysis with Deep Learning Models for Maternal Endpoints.

**eTable 6. Trends in prevalent cardiovascular comorbidities and CVD and incident CV complications in pregnancy from 2001 to 2019 in PADME**

| | Overall 2001-2019 | 2001-2005 | 2006-2010 | 2011-2015 | 2016-2019 | Trend | |
|---|---|---|---|---|---|---|---|
| | | | | | | β (SE) | p-value |
| Pregnancies | 57,683 | 13,715 | 16,495 | 16,516 | 10,957 | - | - |
| Maternal age range, years | 28 - 38 | 26 - 38 | 26 - 38 | 28 - 38 | 29 - 39 | - | - |
| **Prevalent maternal cardiovascular comorbidities and CVD** | | | | | | | |
| Pre-existing obesity, n (%) | 7068 (12%) | 836 (6%) | 1800 (11%) | 2478 (15%) | 1954 (18%) | 0.072 (0.002) | <0.0001 |
| Pre-existing hypertension, n (%) | 4486 (8%) | 763 (6%) | 1154 (7%) | 1450 (9%) | 1119 (10%) | 0.034 (0.003) | <0.0001 |
| Pre-existing hyperlipidemia, n (%) | 5953 (10%) | 1074 (8%) | 1857 (11%) | 1830 (11%) | 1192 (11%) | 0.008 (0.003) | 0.002 |
| Pre-existing diabetes, n (%) | 1786 (3%) | 347 (3%) | 520 (3%) | 544 (3%) | 375 (3%) | 0.014 (0.005) | 0.002 |
| Pre-existing CVD, n (%) | 2394 (4%) | 302 (2%) | 672 (4%) | 775 (5%) | 645 (6%) | 0.050 (0.004) | <0.0001 |
| **Incident pregnancy-related CV complications within 1 year of delivery** | | | | | | | |
| Overall CV complications, n (%) | 8457 (15%) | 1828 (13%) | 2286 (14%) | 2362 (14%) | 1981(18%) | 0.015 (0.002) | <0.0001 |
| Maternal death, n (%) | 7 (0.01%) | 1 (0.01%) | 1 (0.01%) | 3 (0.02%) | 2 (0.02%) | - | - |
| MACE, n (%) | 2198 (4%) | 481 (4%) | 643 (4%) | 658 (4%) | 416 (4%) | 0.001 (0.004) | 0.79 |
| HDP, n (%) | 6678 (12%) | 1424 (10%) | 1749 (11%) | 1837 (11%) | 1668 (15%) | 0.020 (0.002) | <0.0001 |

Maternal age is reported as mean ± SD. Prevalence or incidence rates are reported as %. Values shown exclude missing data. Abbreviations: CV = cardiovascular, CVD = cardiovascular disease, HDP = hypertensive disorders of pregnancy, MACE = major adverse cardiovascular event

**eTable 7. Trends in annual age-adjusted prevalent cardiovascular comorbidities and CVD and incident CV complications in pregnancy from 2001 to 2019 in PADME**

| | Overall | 2001 | 2002 | 2003 | 2004 | 2005 | 2006 | 2007 | 2008 | 2009 |
|---|---|---|---|---|---|---|---|---|---|---|
| Pregnancies | 57683 | 2476 | 2653 | 2805 | 2903 | 2878 | 3054 | 3138 | 3452 | 3491 |
| Maternal age range, years | 28 - 38 | 25 - 37 | 26 - 38 | 26 - 38 | 26 - 38 | 26 - 38 | 26 - 38 | 26 - 38 | 26 - 38 | 26 - 38 |
| Pre-existing obesity, % | 13 | 2 | 4 | 4 | 4 | 6 | 6 | 7 | 7 | 7 |
| Pre-existing diabetes, % | 8 | 1 | 2 | 3 | 2 | 2 | 3 | 2 | 5 | 2 |
| Pre-existing HTN, % | 14 | 3 | 4 | 4 | 6 | 4 | 5 | 5 | 8 | 6 |
| Pre-existing HLD, % | 17 | 3 | 6 | 6 | 9 | 6 | 7 | 13 | 10 | 8 |
| Pre-existing CVD, % | 7 | 1 | 1 | 2 | 1 | 2 | 2 | 2 | 4 | 4 |
| CV complications, % | 17 | 11 | 8 | 13 | 13 | 14 | 13 | 12 | 14 | 16 |
| MACE, % | 5 | 1 | 2 | 4 | 6 | 3 | 5 | 4 | 4 | 2 |
| HDP, % | 13 | 10 | 7 | 9 | 11 | 11 | 8 | 9 | 11 | 14 |
| | **2010** | **2011** | **2012** | **2013** | **2014** | **2015** | **2016** | **2017** | **2018** | **2019** |
| Pregnancies | 3360 | 3506 | 3325 | 3328 | 3171 | 3186 | 3133 | 2995 | 2539 | 2290 |
| Maternal age range, years | 26 - 38 | 27 - 38 | 28 - 38 | 28 - 38 | 28 - 38 | 28 - 38 | 29 - 39 | 29 - 39 | 29 - 39 | 29 - 39 |
| Pre-existing obesity, % | 14 | 16 | 12 | 10 | 15 | 12 | 13 | 13 | 15 | 15 |
| Pre-existing diabetes, % | 4 | 1 | 3 | 3 | 7 | 8 | 7 | 2 | 4 | 3 |

| | | | | | | | | | | |
|---|---|---|---|---|---|---|---|---|---|---|
| Pre-existing HTN, % | 10 | 13 | 9 | 8 | 8 | 8 | 12 | 9 | 11 | 12 |
| Pre-existing HLD, % | 10 | 9 | 12 | 9 | 12 | 13 | 15 | 11 | 11 | 10 |
| Pre-existing CVD, % | 4 | 3 | 4 | 4 | 7 | 12 | 9 | 10 | 9 | 7 |
| CV complications, % | 13 | 17 | 13 | 14 | 15 | 19 | 16 | 12 | 18 | 14 |
| MACE, % | 3 | 2 | 2 | 4 | 6 | 4 | 4 | 2 | 5 | 2 |
| HDP, % | 9 | 15 | 11 | 12 | 10 | 14 | 12 | 10 | 14 | 12 |

Maternal age is reported as mean ± SD. Age-adjusted prevalence or incidence rates are reported as %. Values shown exclude missing data. Abbreviations: CV = cardiovascular, CVD = cardiovascular disease, HDP = hypertensive disorders of pregnancy, HTN = hypertension, HLD = hyperlipidemia, MACE = major adverse cardiovascular event

**eTable 8. Trends in prevalent cardiovascular comorbidities and CVD and incident CV complications in pregnancy from 2001 to 2019 among first pregnancies available in PADME**

| | Overall 2001-2019 | 2001-2005 | 2006-2010 | 2011-2015 | 2016-2019 | Trend | |
|---|---|---|---|---|---|---|---|
| | | | | | | β (SE) | p-value |
| Pregnancies | 38,997 | 11637 | 11125 | 10235 | 6000 | - | - |
| Maternal age range, years | 26 - 38 | 25 - 37 | 26 - 38 | 27 - 38 | 29 - 39 | - | - |
| **Prevalent maternal cardiovascular comorbidities and CVD** | | | | | | | |
| Pre-existing obesity, n (%) | 3650 (9%) | 662 (6%) | 1044 (9%) | 1111 (11%) | 833 (14%) | 0.059 (0.003) | <0.0001 |
| Pre-existing hypertension, n (%) | 2648 (7%) | 618 (5%) | 693 (6%) | 777 (8%) | 560 (9%) | 0.030 (0.004) | <0.0001 |
| Pre-existing hyperlipidemia, n (%) | 3467 (9%) | 834 (7%) | 1092 (10%) | 960 (9%) | 581(10%) | 0.010 (0.003) | 0.003 |
| Pre-existing diabetes, n (%) | 978 (2%) | 275 (2%) | 297 (3%) | 248 (2%) | 158 (3%) | 0.001 (0.006) | 0.84 |
| Pre-existing CVD, n (%) | 1333 (3%) | 250 (2%) | 381 (3%) | 415 (4%) | 287 (5%) | 0.046 (0.005) | <0.0001 |
| **Incident pregnancy-related CV complications within 1 year of delivery** | | | | | | | |
| Overall CV complications, n (%) | 6094 (16%) | 1614 (14%) | 1708 (15%) | 1601 (16%) | 1171 (20%) | 0.017 (0.002) | <0.0001 |
| Maternal death, n (%) | 4 (0.01%) | 1 (0.01%) | 1 (0.01%) | 2 (0.02%) | 0 (0%) | - | - |
| MACE, n (%) | 1499 (4%) | 415 (4%) | 468 (4%) | 400 (4%) | 216 (4%) | -0.001 (0.005) | 0.78 |
| HDP, n (%) | 4890 (13%) | 1270 (11%) | 1322 (12%) | 1282 (13%) | 1016 (17%) | 0.023 (0.003) | <0.0001 |

Maternal age is reported as mean ± SD. Prevalence or incidence rates are reported as %. Values shown exclude missing data. Abbreviations: CV = cardiovascular, CVD = cardiovascular disease, HDP = hypertensive disorders of pregnancy, MACE = major adverse cardiovascular event

**eTable 9. Trends in incident CV complications in pregnancy from 2001 to 2019 in PADME**

| | Overall 2001-2019 | 2001-2005 | 2006-2010 | 2011-2015 | 2016-2019 |
|---|---|---|---|---|---|
| **MACE** | | | | | |
| Venous thromboembolism, n (%) | 1123 (2%) | 245 (2%) | 337 (2%) | 370 (2%) | 171 (2%) |
| Pulmonary embolism, n (%) | 221 (0.4%) | 35 (0.3%) | 48 (0.3%) | 74 (0.5%) | 64 (0.6%) |
| Systemic embolism, n (%) | 35 (0.07%) | 10 (0.07%) | 18 (0.1%) | 3 (0.02%) | 4 (0.04%) |
| TIA, n (%) | 288 (0.5%) | 48 (0.4%) | 91 (0.6%) | 75 (0.5%) | 74 (0.7%) |
| Hemorrhagic CVA, n (%) | 60 (0.1%) | 11 (0.08%) | 30 (0.2%) | 12 (0.1%) | 7 (0.06%) |
| Ischemic CVA, n (%) | 118 (0.2%) | 22 (0.2%) | 31 (0.2%) | 29 (0.2%) | 36 (0.3%) |
| Heart failure, n (%) | 362 (1%) | 103 (1%) | 114 (1%) | 110 (1%) | 35 (0.3%) |
| Myocardial infarction, n (%) | 65 (0.1%) | 26 (0.2%) | 20 (0.1%) | 16 (0.1%) | 3 (0.03%) |
| Vascular dissection, n (%) | 31 (0.05%) | 2 (0.01%) | 7 (0.04%) | 6 (0.04%) | 16 (0.2%) |
| Atrial fibrillation, n (%) | 105 (0.2%) | 25 (0.2%) | 27 (0.2%) | 20 (0.1%) | 33 (0.3%) |
| Supraventricular tachycardia, n (%) | 118 (0.2%) | 6 (0.04%) | 5 (0.03%) | 21 (0.1%) | 86 (0.8%) |
| Ventricular tachycardia, n (%) | 164 (0.3%) | 37 (0.3%) | 44 (0.3%) | 38 (0.2%) | 45 (0.4%) |
| Cardiac arrest, n (%) | 24 (0.04%) | 3 (0.02%) | 8 (0.05%) | 8 (0.05%) | 5 (0.05%) |

| HDP | | | | | |
|---|---|---|---|---|---|
| Chronic hypertension, n (%) | 3038 (5%) | 533 (4%) | 759 (5%) | 909 (5%) | 837 (8%) |
| Gestational hypertension, n (%) | 1010 (2%) | 0 (0%) | 1 (0.01%) | 61 (0.4%) | 948 (9%) |
| Preeclampsia, n (%) | 4628 (8%) | 1176 (8%) | 1319 (8%) | 1319 (8%) | 814 (7%) |
| Eclampsia, n (%) | 89 (0.2%) | 26 (0.2%) | 33 (0.2%) | 24(0.2%) | 6 (0.1%) |
| HELLP Syndrome, n (%) | 17 (0.03%) | 0 (0%) | 0 (0%) | 0 (0%) | 17 (0.2%) |

Abbreviations: CVA = cerebrovascular accident, HDP = hypertensive disorder of pregnancy, HELLP = hemolysis elevated liver enzymes low platelets, MACE = major adverse cardiovascular event, TIA = transient ischemic attack

**eTable 10. Prevalent maternal CVD and incident pregnancy-related CV complications in PADME**

| | Prevalent CVD | | | Incident CVD | | |
|---|---|---|---|---|---|---|
| | **Overall PADME (N = 57,683)** | **Incident CV complication (N = 8457)** | **Without CV complication (N = 49,226)** | **Overall PADME (N = 57,683)** | **Prevalent CVD (N=2394)** | **Without prevalent CVD (N=55,289)** |
| Overall CV disease, n (%) | 2394 (4%) | 876 (10%) | 1518 (3%) | 2198 (4%) | 568 (24%) | 1630 (3%) |
| Venous thromboembolism, n (%) | 1068 (2%) | 374 (4%) | 694 (1%) | 1123 (2%) | 218 (10%) | 905 (2%) |
| Pulmonary embolism, n (%) | 263 (0.5%) | 153 (2%) | 110 (0.2%) | 221 (0.4%) | 118 (5%) | 103 (0.2%) |
| Systemic embolism, n (%) | 81 (0.1%) | 44 (0.5%) | 37 (0.1%) | 35 (0.06%) | 14 (0.6%) | 21 (0.04%) |
| TIA, n (%) | 572 (1%) | 233 (3%) | 339 (1%) | 288 (0.5%) | 131 (5%) | 157 (0.3%) |
| Hemorrhagic CVA, n (%) | 230 (0.4%) | 78 (1%) | 152 (0.3%) | 60 (0.1%) | 17 (0.7%) | 43 (0.1%) |
| Ischemic CVA, n (%) | 199 (0.3%) | 119 (1%) | 80 (0.2%) | 118 (0.2%) | 52 (2%) | 66 (0.1%) |
| Heart failure, n (%) | 300 (0.5%) | 119 (1%) | 181 (0.4%) | 362 (0.6%) | 61 (3%) | 301 (0.5%) |
| Myocardial infarction, n (%) | 100 (0.2%) | 52 (0.6%) | 48 (0.1%) | 65 (0.1%) | 19 (1%) | 46 (0.1%) |
| Vascular dissection, n (%) | 33 (0.06%) | 20 (0.2%) | 13 (0.03%) | 31 (0.05%) | 15 (0.6%) | 16 (0.03%) |
| Atrial fibrillation, n (%) | 147 (0.3%) | 84 (1%) | 63 (0.1%) | 105 (0.2%) | 40 (2%) | 65 (0.1%) |
| Supraventricular tachycardia, n (%) | 114 (0.2%) | 48 (0.6%) | 66 (0.1%) | 118 (0.2%) | 39 (2%) | 79 (0.1%) |
| Ventricular tachycardia, n (%) | 238 (0.4%) | 112 (1%) | 126 (0.3%) | 164 (0.3%) | 64 (3%) | 100 (0.2%) |
| Cardiac arrest, n (%) | 17 (0.03%) | 12 (0.1%) | 5 (0.01%) | 24 (0.04%) | 8 (0.3%) | 16 (0.03%) |

| | | | | | | |
|---|---|---|---|---|---|---|
| Overall HDP, n (%) | 419 (0.7%) | 419 (5%) | 0 (0%) | 6678 (12%) | 419 (17%) | 6259 (11%) |
| Chronic hypertension, n (%) | 225 (0.4%) | 225 (3%) | 0 (0%) | 3038 (5%) | 225 (9%) | 2813 (5%) |
| Gestational hypertension, n (%) | 75 (0.1%) | 75 (1%) | 0 (0%) | 1010 (2%) | 75 (3%) | 935 (2%) |
| Pre-eclampsia, n (%) | 269 (0.5%) | 269 (3%) | 0 (0%) | 4628 (8%) | 269 (11%) | 4359 (8%) |
| Eclampsia, n (%) | 6 (0.01%) | 6(0.1%) | 0 (0%) | 89 (0.2%) | 6 (0.3%) | 83 (0.2%) |
| HELLP syndrome, n (%) | 0 (0%) | 0 (0%) | 0 (0%) | 17 (0.03%) | 0 (0%) | 17 (0.03%) |
| Prior pregnancy-related cardiovascular complication*, n (%) | 541 (1%) | 254 (3%) | 287 (0.6%) | 2622 (5%) | 541 (23%) | 2081 (4%) |

Values are mean ± SD, median (Q1, Q3), or n (%). Values shown exclude missing data. Abbreviations: CV = cardiovascular, CVA = cerebrovascular accident, HDP = hypertensive disorder of pregnancy, PADME = Predictive Analysis with Deep Learning Models for Maternal Endpoints. * Prior pregnancy-related cardiovascular complication among 24,007 pregnancies with ≥1 prior pregnancies.

**eFigure 1.** Example of clinical note with gravidity, parity, and estimated gestational age extracted using regular expressions

**File Type: OB Labor Progress Note**
**Identified Gestational age: 35w3d**
**Identified Gravidity: G5**
**Identified Parity: P2113**

INTRODUCTION:
Assumed care of [removed_age] y/o **G5P2113** @**35w3d** undergoing IOL since [removed_date] for BPP 4/8 in the setting of umbilical vein varix per MFM recommendation

PAST HISTORY:
On last exam, pt was 5/70 ballotable

MEDICATIONS:
Pt is now s/p misoprostol x 1 and Pitocin intermittently since admission.

VITALS:
Temperature: [36.4 ?C (97.6 ?F)-37.1 ?C (98.7 ?F)] 36.4 ?C (97.6 ?F). Heart Rate: [76-90] 84 Respiratory Rate: [18] 18 BP: (92-124)/(54-83). 122/78

GENERAL:
NAD FHT: 110s-120s, mod LTV, +accels, no decels Toco: irregular, Pit @ 8 nU/min SVE: Dilation: 5; Effacement (%): 70; Station: Floating Labs: GLU POC Date Value Ref Range Status [removed_date] 66 (L) 70 – 100 mg/dL Final [removed_date] 83 70 – 100 mg/dL Final [removed_date] 91 70 – 100 mg/dL Final [removed_date] 70 70 – 100 mg/dL Final [removed_date] 47 (L) 70 – 100 mg/dL Final [removed_date] 56 (L) 70 – 100 mg/dL Final. GBS cx – pending

ASSESSMENT
A [removed_age] female **G5P2113** at **35w3d**, hx Type II DM, undergoing IOL for BPP 4/8 in the setting of umbilical vein varix, now with reassuring Cat I FHT, GBS unknown Plan: 1. D/w pt need for AROM to augment labor at this point. Given high station, there is a concern for cord prolapse with amniotomy; given this, pt was offered AROM in the OR vs elective primary C/S; risks of both. Pt opts for trial of AROM in OR with emergent C/S only in the setting of prolapse 2. Continue management of labor induction with Pitocin; await better ctx pattern prior to proceeding with AROM 3. Epidural to be redosed prior to heading back to OR 4. Type II DM – overall good glycemic control; will monitor BS hourly and given D5 prn; pt to stay NPO at this time 5. Cont Vanco for unknown GBS; f/u culture 6. New Type + screen sent 7. Work toward vaginal delivery

Depicted is a labor and delivery note with gravidity, parity, and gestational age data extracted via regular expressions depicted at the top and shown in bold within the note text. The note has been edited for brevity and to remove potentially identifying information.

**eFigure 2.** Defining the Pregnancy Episode.

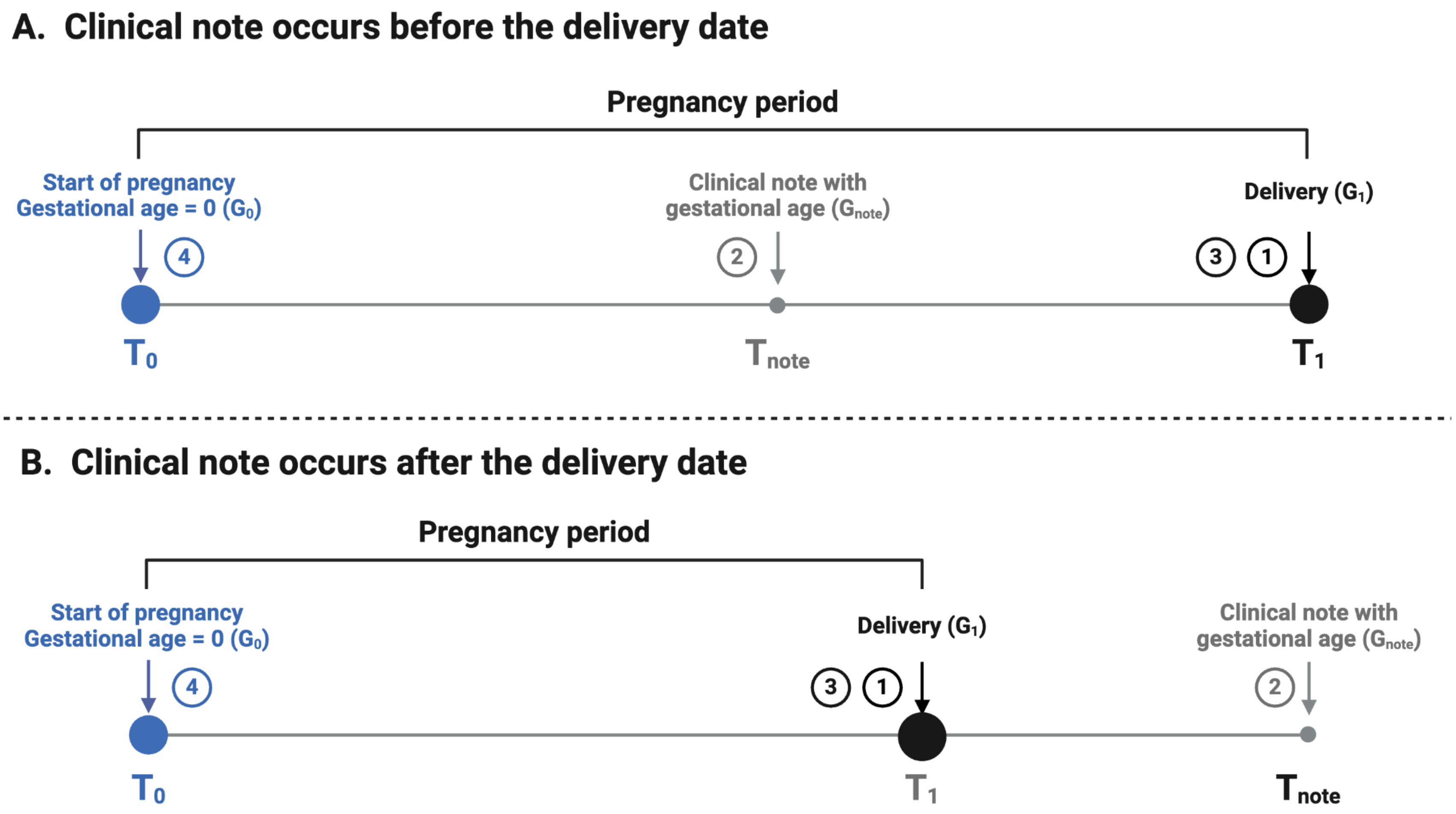

This figure summarizes the stepwise approach used to define the pregnancy episode when the clinical note with documented gestational age occurs before the delivery date (**Panel A**) and after the delivery date (**Panel B**). Abbreviations: $T_0$ = date of start of pregnancy, $T_1$ = date of pregnancy endpoint. $T_{note}$ = date of clinical note with documented gestational age, $G_0$ = gestational age at start of pregnancy. $G_1$ = gestational age at delivery, $G_{note}$ = gestational age documented in clinical note.

**eFigure 3.** Trends in age-adjusted prevalence of maternal cardiovascular comorbidities.

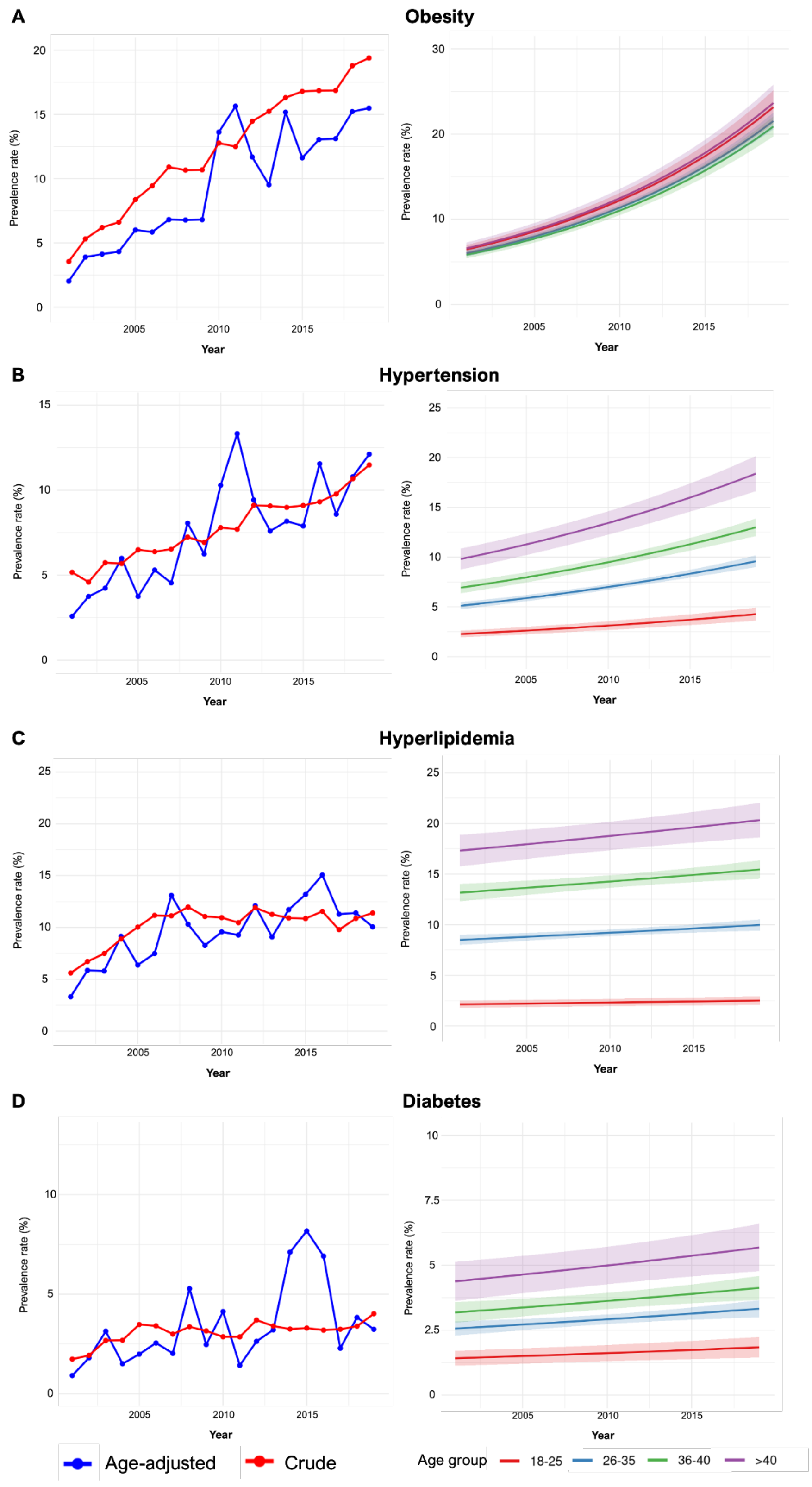

**Left panels** display the crude and age-adjusted annual trend in prevalence of maternal CV comorbidities in pre-specified time epochs from 2001 to 2019. **Right panels** display the annual trend in prevalence of maternal CV comorbidities fitted using the Poisson regression stratified by maternal age group. Cardiovascular comorbidities include obesity (**Panel A**), hypertension (**panel B**), diabetes (**panel C**), and hyperlipidemia (**panel D)**.

**eFigure 4.** Trends in age-adjusted prevalence of pre-existing maternal CVD from 2001 to 2019.

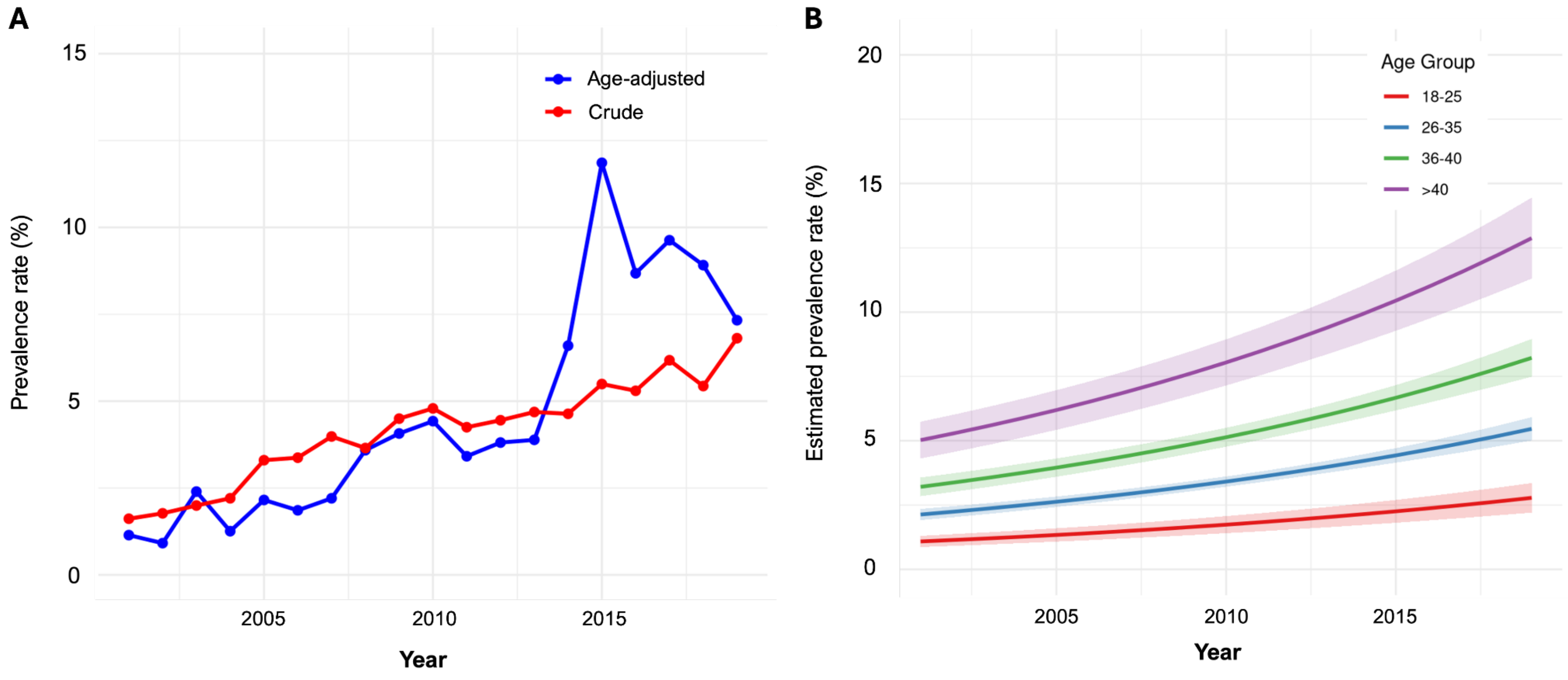

**Panel A** displays the crude and age-adjusted annual trend in prevalent maternal CVD in pre-specified time epochs from 2001 to 2019. **Panel B** displays the annual trend in maternal CVD prevalence using the Poisson regression stratified by maternal age group.

**eFigure 5.** Trends in age-adjusted incidence of pregnancy-related cardiovascular complications.

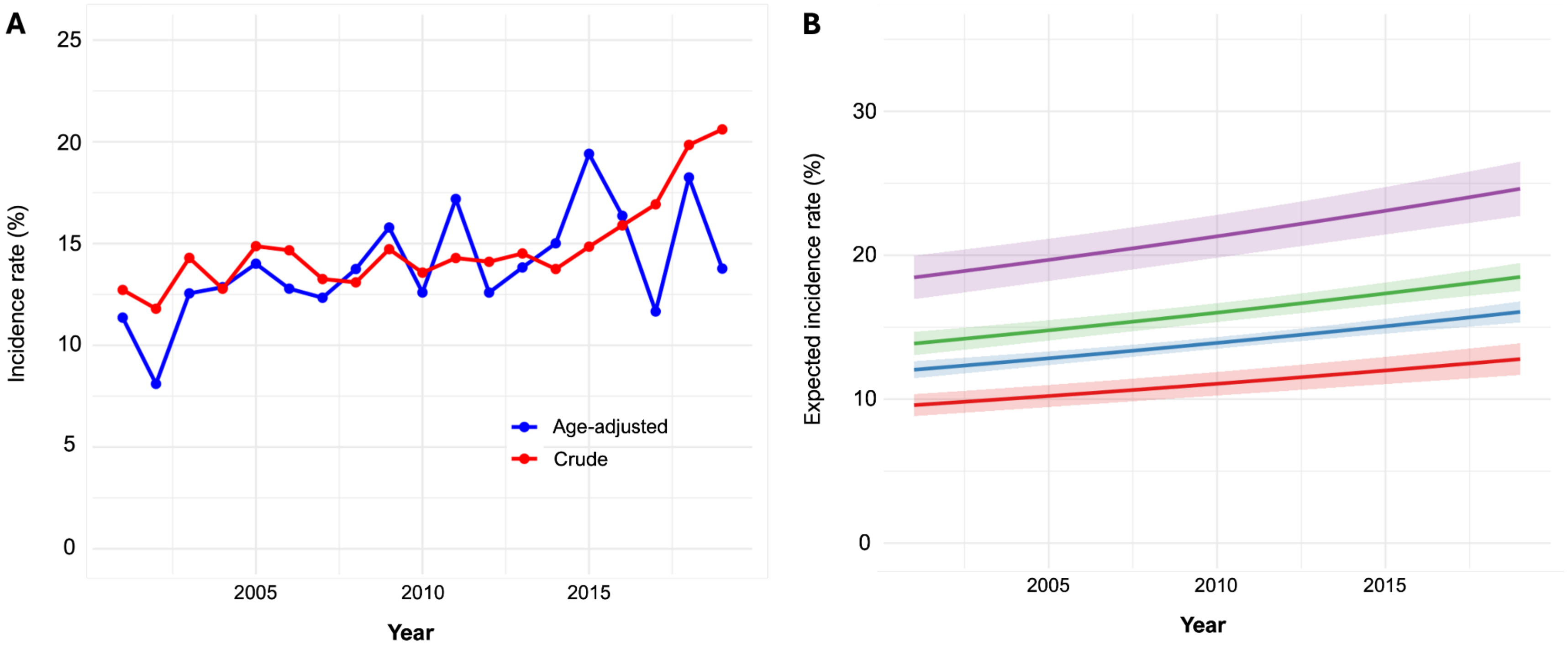

**Panel A** displays the crude and age-adjusted annual trend in incident pregnancy-related CV complications in pre-specified time epochs from 2001 to 2019. **Panel B** displays the trend in incidence of pregnancy-related CV complications fitted using the Poisson regression stratified by maternal age group.
